# New phenotype discovery method by unsupervised deep representation learning empowers genetic association studies of brain imaging

**DOI:** 10.1101/2022.12.10.22283302

**Authors:** Khush Patel, Ziqian Xie, Hao Yuan, Sheikh Muhammad Saiful Islam, Wanheng Zhang, Assaf Gottlieb, Han Chen, Luca Giancardo, Alexander Knaack, Evan Fletcher, Myriam Fornage, Shuiwang Ji, Degui Zhi

## Abstract

Understanding the genetic architecture of brain structure is challenging, partly due to difficulties in designing robust, non-biased descriptors of brain morphology. Until recently, brain measures for genome-wide association studies (GWAS) consisted of traditionally expert-defined or software-derived image-derived phenotypes (IDPs) that are often based on theoretical preconceptions or computed from limited amounts of data. Here, we present an approach to derive brain imaging phenotypes using unsupervised deep representation learning. We train a 3-D convolutional autoencoder model with reconstruction loss on 6,130 UK Biobank (UKBB) participants’ T1 or T2-FLAIR (T2) brain MRIs to create a 128-dimensional representation known as endophenotypes (ENDOs). GWAS of these ENDOs in held-out UKBB subjects (n = 22,962 discovery and n = 12,848/11,717 replication cohorts for T1/T2) identified 658 significant replicated variant-ENDO pairs involving 43 independent loci. Thirteen loci were not reported in earlier T1 and T2 IDP-based UK Biobank GWAS. We developed a perturbation-based decoder interpretation approach to show that these loci are associated with ENDOs mapped to multiple relevant brain regions. Our results established unsupervised deep learning can derive robust, unbiased, heritable, and interpretable endophenotypes from imaging data.

## Main

Structural magnetic resonance imaging (MRI) modalities such as T1 and T2-FLAIR (T2) scans enable the study of brain anatomy and pathology in high resolution. With the availability of large cohorts with both brain MRI and genetic information^1–5^, genome-wide association studies (GWAS) of the brain structures have shed light on the genetic factors underlying the variations in brain morphology and can potentially aid the understanding of etiopathology of neuropsychiatric disorders. Still, one of the main methodological challenges for brain imaging GWAS is to derive comprehensive, heritable, and interpretable representations of the brain from complex 3D brain MRIs. Most existing GWAS studies^6–10^ use phenotype values capturing volumes of brain regions, cortical surface area and cortex thickness estimated by classical software such as FSL^11^, FreeSurfer^12^ and SPM^13^. One of the most extensive efforts of this type was the UK Biobank (UKBB) brain imaging genetics study^14,15^, where an extensive set of 3,144 brain imaging-derived phenotypes (IDPs), directly measurable features derived from brain image by algorithmic processing, from the UKBB was studied and 148 and 692 clusters of association between genetic variants and IDPs were identified respectively. Also, a GWAS of FreeSurfer-derived vertex-based measures using UKBB data identified 780 loci for cortical thickness and surface area^16^.

However, traditional approaches for deriving phenotypes from brain MRI have limitations. Brain-derived IDPs are always subject to ambiguity and uncertainty due to many factors. For example, segmentation of brain into regions of interest (ROIs) was often a prerequisite for downstream processing. However, even widely-used standard region segmentation software still has biases and inconsistencies^17–19^. One such bias is the partial volume effects, i.e., a single image voxel may contain several types of tissues due to the finite spatial resolution of the imaging device^17^. Also, the brain segmentation algorithms based on image registration may be affected significantly by artifacts and pathology^17^.

In addition to issues arising from traditional approaches to segmenting brain IDPs, recent commentaries have pointed out inherent limitations from modeling outcomes associated with single brain measures from pre-selected lists^20^. The essence of this critique is that portions of multiple IDPs may be associated with an outcome of interest (e.g., cognitive performance or genetic association), crossing traditional ROI boundaries to recruit subsets of multiple regions while not using the entirety of any region. Modeling brain-outcome associations with single whole IDPs thus loses both anatomical specificity (by forcibly incorporating more of a single IDP than is really associated to outcome) and sensitivity (failing to incorporate portions of other IDPs that are also associated)^20^. This is true *a fortiori* in GWAS, due to the pleiotropy or distributed influence of a genetic locus (SNP) on multiple brain regions and across different imaging modalities^21^.

In response, some multivariate statistical approaches have modeled SNP associations with multiple brain measures, obtaining increased sensitivity and enhanced loci discovery. However, these approaches still require input of precomputed brain measures in some form. This necessitates *a priori* selections of measures that are expected to be relevant, with the risk of overlooking others. For example, a multivariate association study of cortical vertices^16^ captures some cortical features but omits subcortical features that could also discover important brain-genome relations.

In the general MRI imaging domain, deep learning (DL) methods, especially convolutional neural networks (CNNs), have demonstrated their power to learn useful features for predicting brain-related phenotypes^22^. However, there has been minimal success in using deep learning to generate brain imaging endophenotypes for GWAS. One of the misconceptions is that extensive labeling is needed to train a DL model for phenotyping MRIs.

Here, we propose unsupervised learning for deriving phenotypes from brain MRIs. Our deep learning approach has the potential to address sensitivity and specificity issues without requiring a precomputed set of brain measures. Specifically, we trained 3D convolutional autoencoders with reconstruction loss and used the bottleneck layer as a vector representation for the input MRI. This unsupervised approach computes a set of “latent” brain measures that implicitly combine features of the whole brain image to best encode individual brains in large training sets. The only criterion in the training is the ability to reproduce an image from its encoding. Since the encoding is a data reduction, there will be loss, but this is determined by the architecture of the neural net rather than an a priori judgment as to which brain features are relevant. Therefore, our approach has the potential to go beyond a priori measurements to generate measures with improved power for genetic discovery.

In this study, we trained our model on UKBB’s T1 and T2 MRIs. The extracted features in the bottleneck layer were then used as endophenotypes (ENDOs) for GWAS. Using the decoder as the generator, a perturbation-based decoder interpretation (PerDI) approach was developed to map the ENDOs to brain regions. These visual associations were corroborated by results from prior research. Our results suggest that this novel, label-free approach can be used to derive interpretable and heritable endophenotypes, empowering the discovery of the genetic architecture of the brain.

## Results

### Overview

The overall rationale of the study is to leverage the large sample size of UKBB brain imaging data to train an unsupervised deep learning model to generate endophenotypes and then conduct genetic association studies over UKBB data with both imaging and genetics data. Our overall analysis framework can be divided into 4 phases: data set selection, deep learning model development, GWAS, and interpretable deep learning model for mapping endophenotypes to brain regions (Figure 1). Details of these phases are provided in the Methods section.

**Figure 1.**
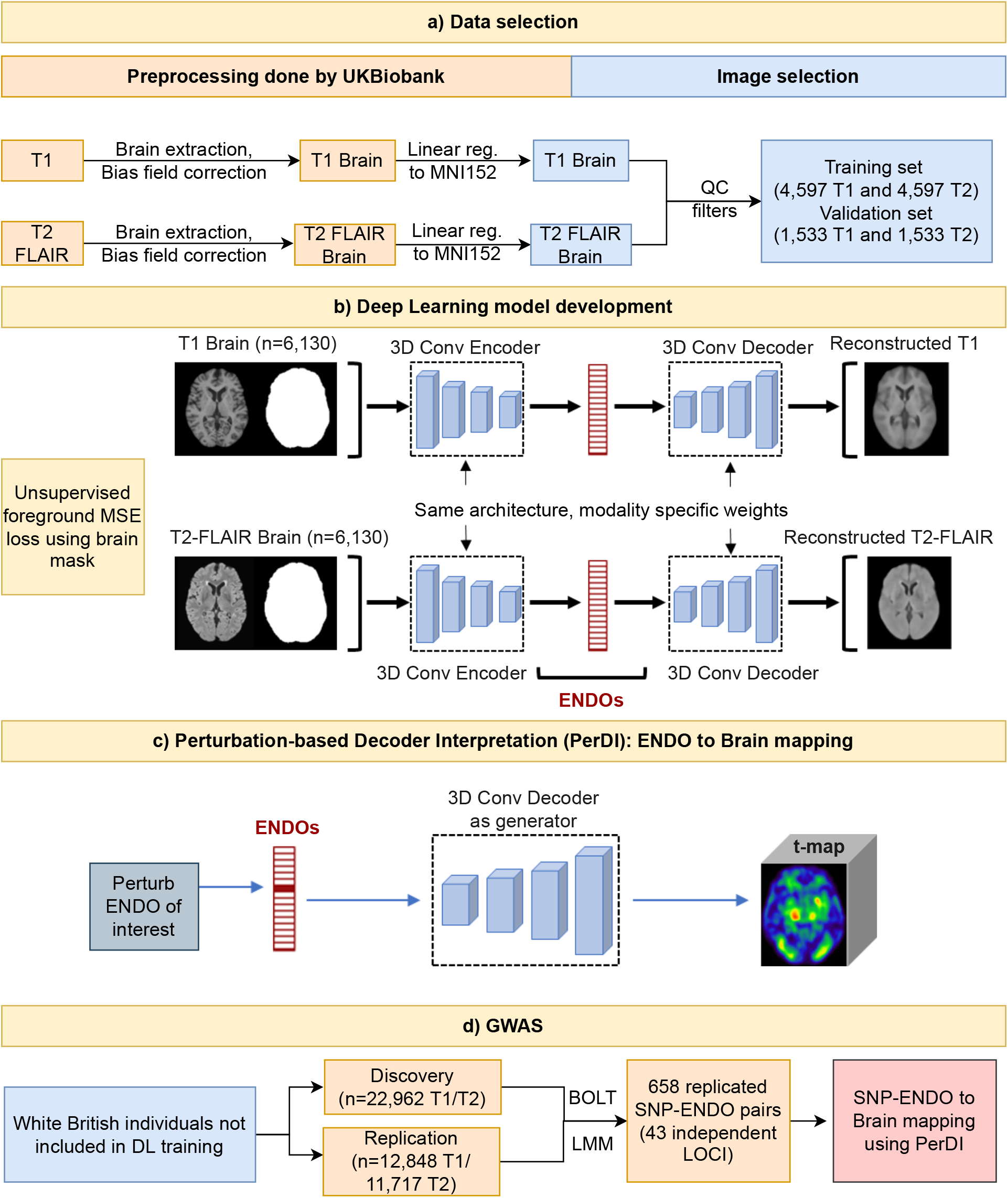
Overall pipeline of the study. a) T1 and T2 FLAIR brain MRI preprocessed by UKBB were divided into separate datasets for deep learning model development and conducting GWAS. b) The autoencoder architecture was trained by background masked mean square error (MSE) loss. c) Perturbation-based Decoder Interpretation (PerDI) method was developed to map ENDOs to brain regions. d) GWAS conducted using a discovery and replication set identified and replicated 658 SNP-ENDO pairs organized into 43 loci. PerDI was used to map SNP-ENDO pairs to brain regions.

A total of 46,099 T1 (44,181 subjects) and 45,294 T2 (43,381 subjects) UKBB MRIs were downloaded. To avoid data leakage, we use disjoint data sets for model development and genetic association study. A dataset of 6,130 images from subjects of mixed ethnicities was chosen as the model development set. This diverse dataset enables the model to learn a greater coverage of variabilities in brain morphology. GWAS was carried out on predictions generated by the model on a separate dataset of White British subjects consisting of 37,376 T1 weighted images and 36,231 T2 weighted images not included in the model development phase. GWAS was performed by dividing the subjects into discovery (22,962 T1 and 22,962 T2) and replication groups (12,848 T1 and 11,717 T2). A detailed data set selection process is shown in Supplementary Table 1.

To derive a compact representation of the input brain image, we use a 3D convolutional autoencoder. Autoencoder is a general architecture for deriving compact representations of any type of input object^23^. For 2D or 3D images, autoencoders with convolutional neural networks (CNN) architectures are a natural choice. While our architecture has semblance of the well-known U-net^24^, we do not introduce the skip connections between the encoder blocks and decoder blocks as we aim to retain maximal information through the bottleneck layer instead of generating sharp images at the high resolution. Model checkpoints with the lowest validation loss were used. The training is effective as we see the difference between the original and the reconstructed images are much lower than that of random pairs (Supplementary Figure 1c).

Also, it is reassuring to see the reconstruction loss in the test set (GWAS set) is similar to that of the validation set even though they are from different ethnicities (Supplementary Figure 1a and 1.b). Although obtaining a high-quality reconstruction is not our primary goal, a visual inspection (see Supplementary Figure 2 for examples of reconstructed images and Supplementary Figure 3-4 for lightbox views of original and reconstructed images) revealed that the reconstructed images share the general shape and anatomy of the original ones. However, many high-resolution features are not reconstructed due to the lack of skip connections to guarantee optimum data retention in ENDOs.

We performed single nucleotide polymorphism GWAS for each ENDO as a phenotype using linear mixed models. We identified 1,133 significant SNP-ENDO pairs with 271 significant SNPs in the discovery cohort (p<5×10^−8^/256). A total of 154 SNPs organized into 43 independent loci were replicated (p<0.05/1133). We identified 13 loci not found in traditional T1/T2 IDP GWAS indicating the power of our approach for phenotype discovery^14,15^.

### Characterization of the endophenotypes

Both the 128-dimensional vectors for T1 and T2 are multivariate representations of the content of the input image. As we do not induce any structures among the 128 dimensions, we expect individual dimensions to be orderless and interchangeable. Indeed, we observed that all 256 dimensions are unimodal and normally distributed (Supplementary Figure 5). Interestingly, there is a lack of general correlation (Supplementary Figure 6) or subcluster structures (Supplementary Figure 7) among them. This lack of correlation is not by design, but it indicates that from the totality of possible brain image information we have found 128 uncorrelated, independent dimensions. Also interesting is that the average absolute correlation within T1 ENDOs and T2 ENDOs (0.1031 and 0.1052, respectively) is smaller than that across T1 and T2 ENDOs (0.1128). This indicates that T1 and T2 embeddings each capture within-modality uncorrelated structural features while additionally capturing some overall features about the brain anatomy.

While it is challenging to interpret the 128-dimensional ENDOs directly, we verify that they capture relevant information. Using multiple linear regression analyses, ENDOs can accurately predict participants’ sex with the area under the ROC curve (AUROC) value for T1 of 0.9840 (0.0021) and T2 of 0.9781 (0.0001). This performance is comparable with existing literature for direct CNN-based sex prediction: 0.95 AUC using T1 and 0.92 AUC using T2^25^, 0.92 accuracy using T1 MRI^26^, 0.80 accuracy using T1^27^,and 0.99 accuracy using T1^28^. Also, age can be predicted from ENDOs with mean absolute error of 3.3664 (0.0705) years for T1 and 3.1249 (0.0439) years for T2. Again, this performance is comparable to or better than existing CNN-based methods using T1 and T2 to predict age: MAE: 4.006 using T1 MRI^29^, MAE: 2.97 to 3.96 years using T2^30^, MAE: 2.14 years using T1 MRI^28^.

Compared with the traditional IDPs, the ENDOs are uniquely more informative: The ENDOs capture the overall shape and brain anatomy through reconstruction, which IDPs cannot. Still, we use multiple linear regressions to understand the linear correlations among the ENDOs and the FSL-derived volume IDPs. For predicting volumes of brain regions from ENDOs, the highest values of coefficient of determination (R^2^: T1/T2) were seen with volume of the following regions: ventricular cerebrospinal fluid (CSF) (0.9849/0.9625). This is due to the fact that in a typical T1/T2-FLAIR MRI, the ventricular CSF is the largest dark region inside the brain mask with a clear boundary, which is best captured by the MSE loss we used. Other regions of high prediction R^2^ are peripheral cortical gray matter (0.6993/0.6925), gray + white matter (0.7384/0.7242), gray matter (0.6928/0.6805), and thalamus (0.6664/0.6093) for both T1 and T2. On the other hand, for predicting the ENDOs from the volume IDPs, the highest R^2^ are about 0.568 for both T1 and T2 and the overall R^2^ values are not high. Complete list of R^2^ values can be found in Supplementary Table 2.

To visualize the population distribution of multivariate ENDOs, we used Uniform Manifold Approximation for unsupervised dimension reduction (UMAP)^31^ to reduce the 128-dimensional ENDOs into 2D (Figure 2 and Supplementary Figures 10-12). For UMAP, most participants’ ENDOs are distributed within a large continuous region, except some small groups of participants (24 T1 ENDOs, 18 T2 ENDOs) have ENDOs in isolated islands, probably due to having high volume of subcortical structures (Figure 2, Supplementary Table 3). The ENDOs are clearly correlated with volume of ventricular CSF, consistent with the multiple regression analyses. The correlations of ENDOs with age and sex are also visible (Supplementary Figure 10). The components derived from UMAP for T1 and T2 ENDOs are correlated (Component 1 T1/T2: *r*=0.38, Component 2 T1/T2: *r*=0.77) (Supplementary Table 4). In addition, principal component analysis (PCA) (Supplement Figure 8) and t-distributed stochastic neighbor embedding (tSNE) (Supplement Figure 9) analyses of ENDOs show a similar separation of groups as UMAP. Of note, t-SNE seems to be capturing more local patterns. The reconstruction shows the shape, volume, texture, and anatomic relationship captured by the ENDOs.

**Figure 2.**
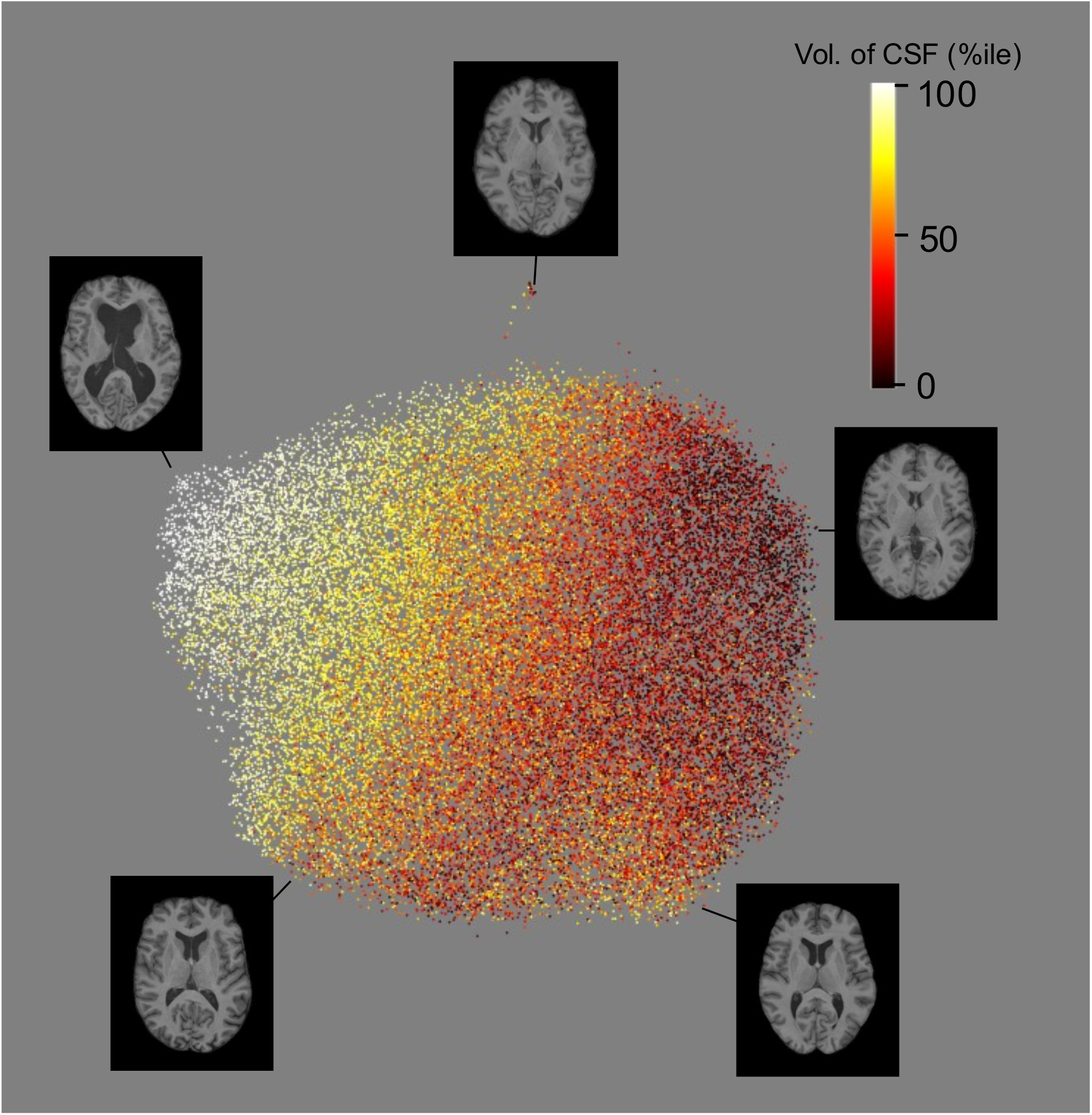
UMAP visualization of T1 ENDOs demonstrated their correlation with brain volume measures. ENDOs of 37,376 T1 are reduced into two components using UMAP and colored with volume of ventricular CSF ranked as percentile. Axial slices of the T1 brain show variation in CSF as colored by UMAP and demonstrate the patterns captured by the ENDOs. x- and y-axes are arbitrary up to translations and rotations.

### Interpretable model for mapping endophenotypes to brain regions

To identify relevant brain regions for our ENDOs, we design and adopt a perturbation-based Decoder Interpretation (PerDI). This is because we are mainly concerned about how the changes to our ENDOs translate to the variability of brain MRI images. Briefly, for an ENDO dimension of interest, we generate perturbed reconstructions by adding one standard deviation to the dimension of interest of the ENDO vector of an input image. We use voxel-wise paired t-tests to compare the original and the perturbed and original reconstructions for 500 randomly selected brain images. For each ENDO, we generate a smoothed t-map to highlight its most relevant brain regions. See Methods: Decoder interpretation for detailed description of our methods.

Although no brain segmentation and region annotation were used during training, some ENDOs hit on punctuated subcortical structures as quantified by the enrichment of high-ranking voxels in regions of interest (Figure 3). We use brain structures in the Harvard-Oxford cortical and subcortical structural atlas to annotate prominent regions in t-map and use Kolmogorov-Smirnov (K-S) statistics to quantify the match of the brain structures (see Methods: t-map annotation). In general, the t-map of ENDOs are often not coincident to ROIs, as they are derived in an unsupervised fashion. We typically see single ENDOs can span multiple atlas-defined brain structures with different weights. For example, ENDO 64 of T2 (T2:64) is found to represent multiple regions in the frontal lobe and lateral ventricles. K-S statistic values for all dimensions can be found in Supplementary Table 5.

**Figure 3.**
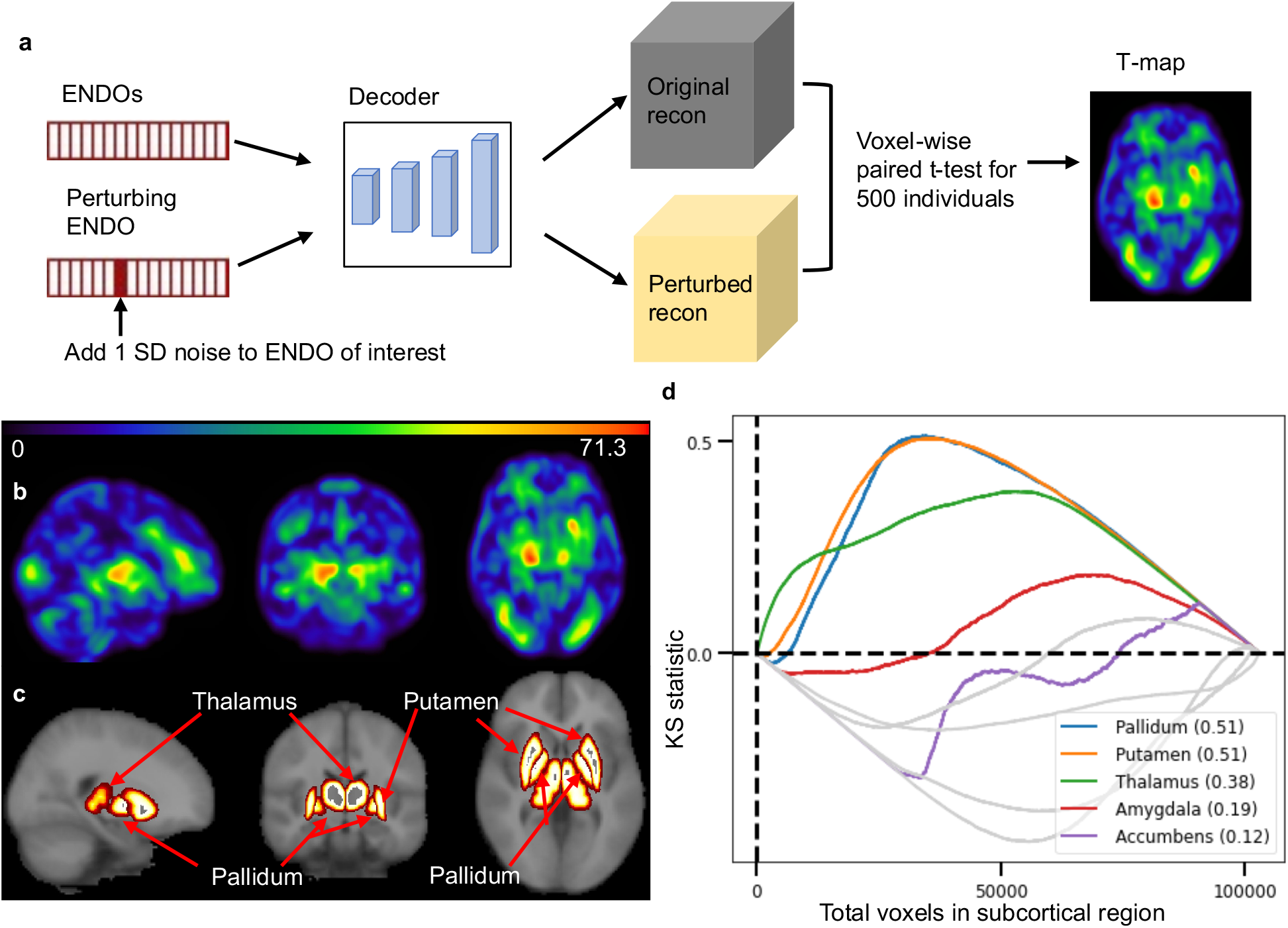
Perturbation-based decoder interpretation (PerDI). a) One standard deviation is added to the dimension of interest in the ENDOs of one input image to generate a perturbed reconstruction image. The differentials of the perturbed and original reconstruction highlight the voxels relevant to the dimension of interest. We repeat this for 500 randomly selected images and use paired t-tests to generate a voxel-wise t-map. b) absolute value of t-map generated for T2:67 by PerDI. c) the Harvard-Oxford atlas labels for relevant subcortical structures. d) Kolmogorov–Smirnov statistic is computed to identify subcortical regions of importance in the t-map generated by PerDI. K-S enrichment can highlight small regions that might not be prominently visualized in the t-map. Plot shows putamen (K-S: 0.51), pallidum (K-S: 0.51), and thalamus (K-S: 0.38) as the most prominent regions.

Comparing the regional enrichment between all ENDOs and all brain regions (Supplementary Table 5), the ENDOs have decent coverage of most regions: For T1 and T2, 43 out of 47 cortical regions have some ENDOs with K-S statistic > 0.33 and 7 out of 11 subcortical regions have some endos with K-S statistics > 0.40 respectively. See Supplementary Figures 13-16 for maximum K-S statistics.

### Genetic association study of endophenotypes

We conducted single nucleotide polymorphism (SNP) variant genome-wide association studies (GWASs) for each ENDO as phenotype using linear mixed models over the array-genotyped markers. In the discovery phase, to adjust for multiple testing of a total of 256 ENDOs from both T1 and T2, the p-value threshold is set to (5×10^−8^)/256. We identified 1,133 significant SNP-ENDO pairs with 271 significant SNPs. A total of 154 SNPs organized into 43 independent loci were replicated (p=0.05/1133) (Supplementary Table 6). A total of 43 loci are replicated and listed in Supplementary Table 7 and Extended Data Figure 1. See Methods: Genome-wide association study (GWAS) for detailed description of methods. Overall, the genome inflation factor is well-controlled (Supplementary Figure 17). All loci are close to previous GWAS loci with brain-related phenotypes (p<5×10^−8^) according to the GWAS catalog (see Methods: Querying GWAS catalog). All 43 loci were previously significantly associated (p<5×10^−8^) with structural brain morphology traits, 30 with neurological disease traits, 14 with psychiatric disease traits, and 26 with Alzheimer’s disease-related traits (see Supplementary Table 8).

We anticipate identifying loci that have not been identified by single IDP-based approaches due to the fact that ENDOs are not restricted to single, specific anatomical regions and can extend across multiple regions while encoding image reconstruction information. Notably, our method identifies 13 additional loci (Table 1) that were not previously reported by the Big40 study, which utilized all available conventional image-derived phenotypes (1,437 phenotypes) from T1 and T2-FLAIR brain MRI modalities^14,15^. All these thirteen loci were previously associated (p<5×10^−8^) with brain structure and neurodegenerative disorders in the GWAS catalog.

**Table 1:** ENDO GWAS identified loci previously missed in IDP GWAS. Previous GWAS findings of neurological disease related and brain morphology related traits were annotated through querying GWAS catalog. Top 2 Cortical and subcortical regions relevant to lead ENDO with K-S statistic > 0.2 is shown. Refer to **Methods: Querying GWAS catalog** for more details. *p >5e-8 Abbreviations: BM (brain measurement), CSA (cortical surface area), CT (cortical thickness), LV (Lateral ventricle), C. (Cortex), G. (Gyrus), WMH (White matter hyperintensity), ant (anterior), mid (middle), post (posterior), sup (superior), post-op (post-operative), MS (multiple sclerosis), AD (Alzheimer’s disease),

|  |  |  | Traits associated with locus by prior GWAS |  | Relevant brain regions of lead ENDO annotated by perturbation-based tmap (top 2) |  |
| --- | --- | --- | --- | --- | --- | --- |
| Lead SNP | Lead ENDO (MRI: DIM) | P val (discovery) | Brain morphology | Neurological disorders | Cortical | Subcortical |
| rs760077 | T1:32 | 4.1E-12 | CT, CSA, occipital lobe vol*, neuroimaging measure, brain vol , BM. | Parkinson disease, CCL2*, brain aneurysm, Bell's palsy*, gait , PHF-tau , Lewy body dementia | Temporal Fusiform C (ant division), Juxtapositional Lobule C | Pallidum, Accumbens |
| rs2900976 | T2:14 | 1.4E-15 | CT, CSA , WM microstructure , BM. | late-onset AD* | Juxtapositional Lobule C, Parahippocampal G (post division) | Hippocampus, Brain-Stem |
| rs272889 | T1:33 | 2.1E-11 | CT, brain vol , BM. | brain aneurysm, MS* | Paracingulate G, Planum Polare | Caudate, Thalamus |
| rs9471128 | T2:2 | 8.0E-15 | CT, CSA ,BM. | neurofibrillary tangles* | Sup Temporal G (ant division), mid Temporal G (ant division) | Pallidum, Putamen |
| rs5883403 | T1:78 | 1.9E-10 | CT, CSA , brain vol , BM. | post-op stroke*, PHF-tau | Heschl's G , Insular C | Caudate, Accumbens |
| rs10264106 | T2:87 | 3.1E-13 | LV vol*, hippocampal vol*, BM | AD, beta-amyloid 1-42*, Ischemic stroke*, frontotemporal dementia* | Supracalcarine C, mid Temporal G (ant division) | Pallidum, Brain-Stem |
| rs11523200 | T2:128 | 7.5E-16 | CT, CSA , BM. | neurofibrillary tangles | Sup Temporal G (ant division) Insular C | LV |
| rs1315400 | T2:64 | 1.2E-12 | CT, CSA , WMH*, BM. | Lewy body dementia* | Sup Frontal G, Mid Frontal G | LV |
| rs72815193 | T2:122 | 1.5E-24 | CSA , BM. | PHF-tau | Juxtapositional Lobule C, Frontal Medial C | Pallidum, Thalamus |
| rs11021216 | T1:16 | 6.5E-15 | CSA , BM. | AD*, multiple sclerosis, PHF-tau | Mid Temporal G (ant division), Frontal Orbital C | Amygdala, Accumbens |
| rs1791316 | T1:82 | 5.6E-14 | brain vol, BM | PHF-tau | Inf Temporal G (ant division) Inf Frontal G (pars opercularis) | Pallidum, Hippocampus |
| rs6054759 | T2:61 | 4.0E-12 | paracentral lobule vol*, CT, CSA , lateral orbital frontal C vol* , BM. | AD* | Parietal Operculum C, Heschl's G (H1, H2) | Brain-Stem |
| rs78110303 | T2:93 | 4.8E-15 | CT, CSA , WM microstructure*, brain vol , BM. | neurofibrillary tangles, Ischemic stroke, gait | Planum Polare, Supramarginal G (ant division) | Pallidum, Putamen |

Interestingly, most ENDOs have higher heritability about 0.3044 ± 0.0388 according to LD score regression (LDSC)^32^ (Extended Data Figure 2), than that of the UKBB T1/T2-FLAIR based IDPs 0.1837 (0.02)^15^(Smith et al. Supplementary Table 1) using LDSC.

We also utilized FUMA pipelines for gene annotation and functional enrichment. For T1 and T2 ENDOs, 432 and 353 genes were identified, respectively. Gene set enrichment tests revealed autism spectrum disorder, schizophrenia and other brain related GWAS catalog gene sets predefined at FUMA were significantly enriched. See Supplementary Note 1 for detailed gene-based GWAS catalog analysis using FUMA.

### Meta-analysis

We conducted an inverse-variance weighted fixed-effect meta-analysis of the discovery and the replication GWAS summary statistics using METAL^33^. With the enhanced sample size, a total of 3,485 significant (P<5×10^−8^/256) SNP-ENDO pairs involving 799 SNPs clustered into 170 loci are identified (Extended Data Figure 3). Using the criteria in Methods: Querying Big40 results, we found 72 loci not close to any loci of previous traditional T1 and T2 IDP GWAS^14,15^ (see Supplementary Table 9). Using the criteria in Methods: Querying GWAS catalog, we identified two unique loci previously unrelated to any brain-related trait. A unique locus on chromosome 5 with lead SNP rs6868292 (p = 9.02×10^−11^) in the intron of the PLPP1 gene was previously not associated with any brain-related trait by GWAS. Using the PerDI, the lead ENDO, T2:93, represents planum polare and supramarginal gyrus (ant and post division) as prominent cortical regions and pallidum, putamen, and thalamus as the prominent subcortical region (Extended Data Figure 4). Another unique locus on chromosome 15 with lead SNP rs4776579 (p = 7.41×10^−11^) located in the intron of MYO9A gene was identified by lead ENDO T2:66. T2:66 is associated with structures such as planum temporale, Heschl’s gyrus, and brainstem, as identified by the PerDI method (Extended Data Figure 5).

### Genetic correlation

We conducted a genetic correlation analysis between the results obtained from the meta-analysis (discovery and replication cohort) and published summary statistics for ten brain-related conditions (see Supplementary Figures 18 and 19). Overall, no individual ENDO-trait pair is significant after Bonferroni correction. Interestingly, ischemic stroke showed the most correlated ENDOs (31 ENDOs) with p<0.05 and the max absolute mean genetic correlation value of 0.4069. Amyotrophic lateral sclerosis (0.3807), major depressive disorder (0.3245), and bipolar disorder (0.2626) are other diseases with high max absolute mean genetic correlation values (p < 0.05). Other diseases with a high number of correlated ENDOs (p <0.05) are autism (27 ENDOs), schizophrenia (27 ENDOs), ADHD (17 ENDOs),and bipolar disorder (16 ENDOs). See Methods: Genetic correlation for more details.

### Use ENDOs for interpreting brain GWAS results

Effectively, our ENDOs can be a bridge between genetic variants and relevant brain regions, whereby providing anatomical details of a genetic association signal. For example, we found ENDO T2:67 is associated with rs13107325 (SLC39A8). This SNP was previously found to be associated significantly (p <5×10^−8^) with thalamic volume^34^, nucleus accumbens volume^8^, schizophrenia^35–38^, alcoholism^39–44^, intelligence^45,46^ and other brain morphology related traits (brain volume measurement, neuroimaging measurement, cortical thickness) in GWAS catalog. Association with Parkinson’s disease^47^ was reported with p=7×10^−8^. We also found rs12146713 (mapped to NAUK1 gene) associated with the same ENDO. rs12146713 was previously found to be associated with thalamus volume (medial thalamic nuclei volume)^34^, diffusion MRI derived white matter microstructure and integrity^15,48^, lateral ventricle volume^15^, cortical thickness and surface area, and subcortical structure volume^21^. These findings are consistent with our PerDI interpretation for this ENDO, which identifies putamen, pallidum, thalamus, amygdala, accumbens and hippocampus as the main subcortical structures (Figure 4d). Supplementary Figures 20-21 shows K-S statistic plots for cortical and subcortical structures respectively for T2:67.

**Figure 4.**
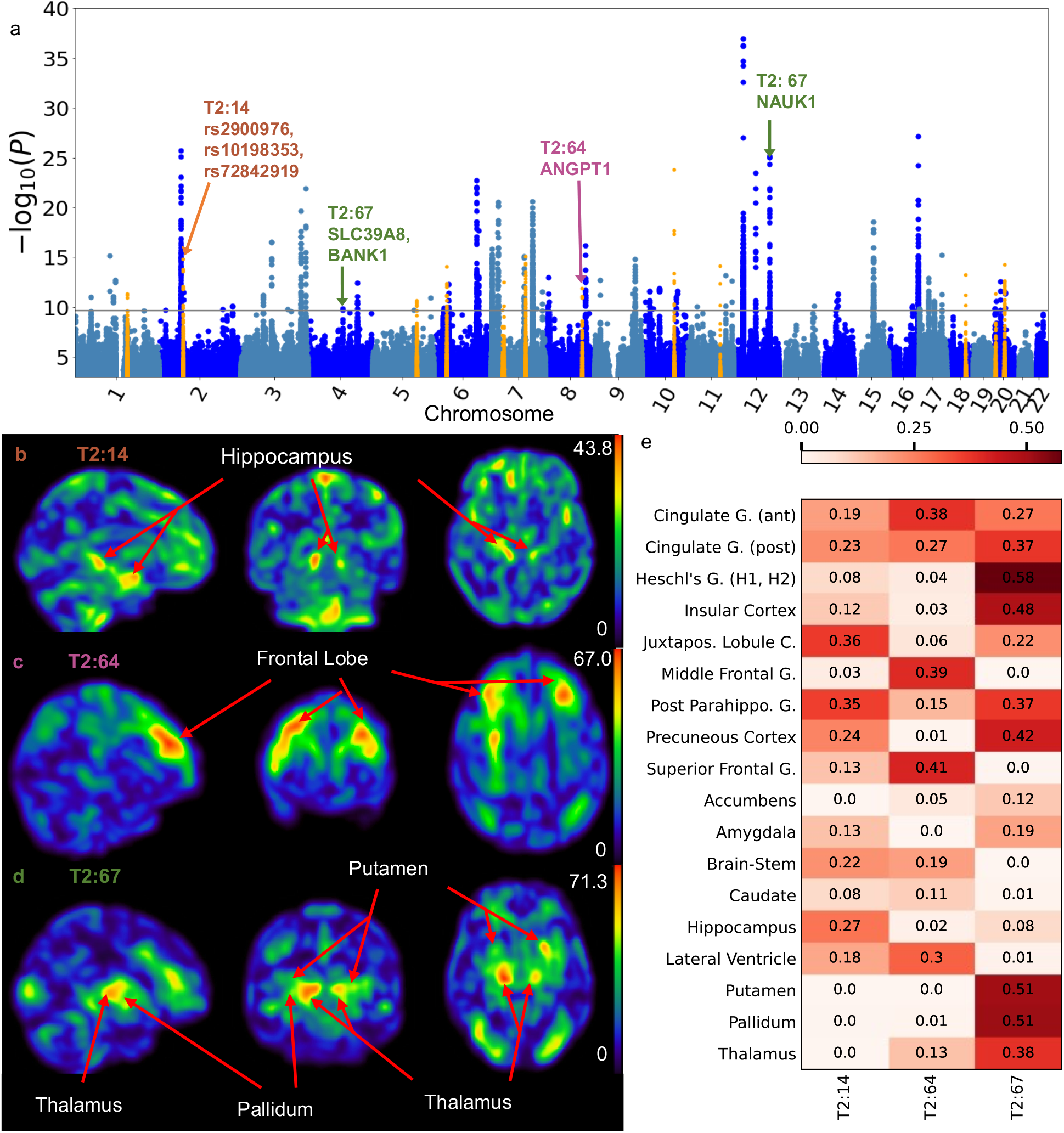
Perturbation-based decoder interpretation (PerDI) to interpret ENDOs associated with specific GWAS loci. a) GWAS discovers and replicates loci identified by ENDOs T2:14, T2:64, T2:67. T2:14 identified locus contains rs72842919 previously associated with Late-onset Alzheimer’s disease, T2:64 identified locus on chr8 containing ANGPT1 gene, T2:67 identified locus containing rs13107325, rs12146713 previously associated with thalamus volume, putamen volume, schizophrenia, and alcoholism. b) T2:14 t-map shows the hippocampus as the prominent region. c) T2:64 t-map shows the frontal lobe as the prominent region. d) T2:67 t-map shows the thalamus, putamen and pallidum as prominent regions. e) K-S statistic values for selected subcortical and cortical regions showing the regions represented by each ENDO.

Of note, this locus was previously implicated to FSL and FreeSurfer derived phenotypes (IDPs) to ventral caudate, putamen, ventral striatum, anterior cingulate cortex and cerebellar regions (Elliott et al., 2018^14^: Extended Data Fig. 1). However, to visualize the MRI regions implicated by this SNP, the gray matter regions of MRI images of carriers and non-carriers were compared, which is not a cheap operation. Instead, the effect of this SNP to relevant brain regions can be efficiently visualized through the ENDO T2:67 (Figure 4d).

#### Acronyms: G (Gyrus), C (Cortex)

One of the locus missed by previous UKBB IDP GWAS was identified by GWAS of ENDO T2:14. The locus was previously found associated significantly (p<5×10^−8^) with cortical thickness, cortical surface area, brain measurement, and white matter microstructure. Locus contained rs72842919 (171 kb to the lead SNP) associated with late-onset Alzheimer’s disease(p=5×10^−7^)^49^. We identified Juxtapositional lobule cortex and parahippocampal gyrus as main cortical regions and hippocampus and brain stem as the main subcortical structures relevant to T2:14 (Figure 4b). Supplementary Figures 22-23 shows K-S statistic plots for cortical and subcortical structures respectively for T2:14.

Another locus missed by previous UKBB IDP GWAS was identified by ENDO T2:64. This locus was previously found to be associated significantly with cortical thickness, cortical surface area, and white matter hyperintensity. In our study, ENDO T2:64 correspond to portions of the frontal lobe (superior frontal gyrus, middle frontal gyrus, cingulate gyrus, frontal pole, precentral gyrus) and lateral ventricle as revealed using PerDI (Figure 4c and 4e). This locus was missed by IDP GWAS possibly because IDPs were only descriptors of individual single regions while this ENDO captures a combination of features from the frontal lobe and the lateral ventricle. Supplementary Figures 24-25 show K-S statistic plots for cortical and subcortical structures respectively for T2:64.

### Comparison of brain volume IDPs and ENDOs

Compared to traditional image-derived phenotypes (IDPs), our unsupervised-deep learning derived ENDOs are distinct in many ways (Table 2).

**Table 2:** Comparison of image-derived phenotypes vs our unsupervised DL-derived endophenotypes.

|  | **IDP (1,437 descriptors of T1/T2 brain regions)** | **ENDOs (128 T1/128 T2 variables)** |
| --- | --- | --- |
| Derived | Software derived | Learned |
| Preprocessing | Heavy | Light |
| Heritability computed using LDSC | 0.18 | 0.304 (T1:0.313, T2: 0.296) |
| Power to reconstruction | No | Yes |
| Redundancy (self-correlation) | High | Low |
| Time required to derive traits | Hours | Seconds |

Primarily, ENDOs are more heritable than IDPs. Individual ENDOs are more heritable than individual IDPs on average. Compounded on the fact that ENDOs are less mutually correlated than IDPs, this indicates that in total ENDOs capture heritable information in the brain more efficiently than IDPs. We postulate the possible reasons for ENDOs’ higher heritability might come from their better minimizing measurement error and reporting error, and ability to capture population variation. ENDOs, unlike IDPs, need minimal image preprocessing and are derived from whole-brain high-quality images without human labels reducing the chance of measurement error and reporting error. Moreover, while IDPs are derived after optimizing loss for each individual separately, ENDOs are derived after training on the entire population using random mini-batches, better capturing necessary total variation in the population to reduce loss. As a result, ENDOs have more flexible definitions of regions, could be features capturing concerted changes combining traits from multiple regions, or only capturing changes in part of a region that is not present in other parts of the same region unlike IDPs that are limited to atlas-defined areas, allowing them to capture unique traits missed by IDPs.

Moreover, what makes ENDOs distinctive from IDPs is that rather than being passive descriptors, our ENDOs are active “predictive encoding” of the input image. Via the decoder, ENDOs allow us to reconstruct, albeit imperfectly, the original input. Although PCA or NMF are also optimizing some sort of reconstruction loss, their quality of reconstruction is not on par with ENDOs.

For training, rather than being generated via a feature-engineering process, our ENDOs are derived from feature-learning. In terms of efficiency, while IDPs are derived with heavy processing that takes hours, ENDOs are derived with minimal preprocessing and take seconds (on GPU though). When trained model weights are shared, our results should be relatively easy to replicate.

## Discussion

We presented an unsupervised deep learning-based approach to capture complex patterns of the brain from the MRI to define endophenotypes (ENDOs) for genetic association studies. Our 3D convolutional autoencoder neural network model was trained by a reconstruction loss function on 6,130 full-sized T1 and T2-FLAIR (T2) weighted brain MRIs from UKBB. Using White British individuals not included in the deep learning training as the test set, we show that the 128 neurons of the bottleneck layer of the autoencoder as endophenotypes can capture the shape and structure of the brain: analysis of ENDOs using encoder-based reconstruction, unsupervised dimension reduction techniques such as UMAP, PCA, tSNE, and statistical methods such as regression showed that a vast amount of information in the input MRI image is captured, including brain’s volume, shape, texture, anatomy, and pathology. More interestingly, these ENDOs are more heritable than most traditional image-derived phenotypes (IDPs). GWAS of ENDOs identified 43 replicated loci, all of them previously associated with brain phenotypes, establishing the validity of our approach.

As a new phenotype discovery approach, our ENDOs are patterns derived from a data-driven approach and capture concerted changes across the entire brain that frequently occur in the population. These patterns go beyond the pre-defined single region volumetric features such as IDPs. We show some ENDOs have good correlation with some anatomical structures and thus are more easily interpretable. However, many ENDOs are capturing more complex patterns and interpretation of them will be future work.

Of note, recognizing the limitations of traditional single IDP-based approaches, post-hoc statistical approaches have been developed to extend single IDP GWASs to multi-IDP GWAS. Specifically, the MOSTest^21^ explicitly modeled “multivariate omnibus” brain measures associated with single SNP via multivariate statistical tests, yielding enhanced loci discovery. However, this method still requires the pre-selection and pre-computation of input variables, necessitating decisions about what to include and what to leave out, and incurring algorithmic limitations, like segmentation, inherent in image preprocessing. For example, the recent application of MOSTest to a set of 1284 cortical surface vertices^16^, was limited to polygonal representations inherent in FreeSurfer, with all the ambiguities of computing a surface mesh, while failing to account for subcortical brain-genome associations. In other words, multivariate association of IDPs may attain more sensitivity than univariate GWAS by modeling multivariate combinations of the measures provided, but the power of such methods still depends on the quality of the individual measures. In a sense, our ENDOs can be viewed as a dictionary of nonlinear brain patterns, automatically and implicitly acquired by deep learning. These could themselves serve as input to multivariate association methods, with the vector of ENDOs as analogs of the array of IDPs in MOSTest, although we have not pursued this approach here because our main focus was phenotype discovery. However, the CNN-learned encoding implicitly extracts related visible brain features, analogous to IDPs, into each ENDO, without the need for selection or the inherent uncertainty of traditional algorithmic processing. This suggests that univariate GWAS with ENDOs is already a step ahead of IDP-based univariate GWAS and may also offer some advantages over multivariate methods that input pre-selected arrays of IDPs.

There are some limitations to our approach. First, the loss metric used for training our model, mean square error (MSE), smoothens the reconstructed images and loses the high-frequency signal. Moreover, higher contrast regions like ventricles tend to be better captured than inner regions with low contrast, such as white matter. Second, MRIs are linearly registered to preserve the overall shape of the brain. However, this has the consequence that the voxels across images are not perfectly aligned, which will affect the correspondence of ENDOs across individuals. We could study the effect of registration in future studies. Third, we used 128-dimensional ENDOs as a tradeoff between compact representation and image information stored in the vector. We could have used a higher number of ENDOs as the 128-dimensional ENDOs were not correlated. Future studies will be conducted to identify the ideal size of the ENDOs. Finally, only internal validation using UKBB data is conducted. Future external validation in a different data set is warranted.

In conclusion, this work represents the first proof-of-concept of using an unsupervised deep learning approach as an imaging phenotyper for GWAS. We believe our unsupervised deep learning-based method can be used to automatically derive and interpret phenotypes from any imaging modality with minimum human intervention.

## Methods

### Detailed dataset descriptions

The UKBB is chosen because it is the largest public brain imaging study and its data were uniformly processed with a standard pipeline. The UKBB data was accessed via approved project 24247. UKBB MRIs were downloaded on October 15, 2021. UKBB^1^ has provided a bias-field-corrected version of the brain-extracted T1-weighted (T1) and T2-weighted FLAIR (T2) images captured mainly using standard Siemens Skyra 3T running VD13A SP4 (as of October 2015), with a standard Siemens 32-channel RF receive head coil. Resolution of T1 is 1×1×1 mm, and resolution of T2 is 1.05×1×1 mm (https://biobank.ctsu.ox.ac.uk/crystal/crystal/docs/brain_mri.pdf). To maximize the generalizability and minimize feature engineering, we followed a simple processing pipeline developed by the UKBB MRI team. UKBB mainly uses FSL to process brain MRIs (https://www.fmrib.ox.ac.uk/ukbiobank/). The main preprocessing done by UKBB involves defacing MRI, brain extraction (using FSL’s BET, and linear and non-linear registration to standard space using FSL FLIRT and FNIRT), and bias field correction (FSL FAST). Bias field corrected brain-extracted T1 and T2 are selected. All MRIs were linearly registered (affine registration with 12 DOF) to standard MNI152 space using the UKBB-provided pre-computed transformation matrix with FSL FLIRT^50^. Linearly registered MRIs were used to ensure the normalization of head sizes and the overall alignment of MRIs between different subjects while also preserving the structural deformation information in the MRI, unlike non-linear registration. Linearly registered, defaced, bias field corrected brain MRIs were used for all our analysis. Further, each affine registered MRI’s intensity was normalized using Z-score normalization by subtracting mean intensity and dividing the result by the standard deviation of the intensity. Background intensity was excluded in computing the mean and standard deviation of the MRI to prevent skewing toward zero. Background comprises the majority of the voxels and is later excluded in the loss calculation.

UKBB has also provided precomputed quality metrics for MRI like UKBB Data field 25735 “inverted contrast-to-noise ratio” and UKBB Data field 25736 “Discrepancy between T2 FLAIR brain image and T1 brain image,” which were used to ensure the quality of the deep learning training set. Lower values in both metrics indicate higher quality. Images with values below 95 percentile for both the quality metrics were selected for deep learning training and validation set. Only one visit was kept if multiple visits were found to ensure uniformity in the dataset. A dataset of 6,130 images was selected consisting of subjects of mixed ethnicities. The dataset was randomly divided into a training set of size 4,597 images (75%) and a validation set of size 1,533 images (25%). See Figure 1 for the overall pipeline of the study. The validation set was used for tuning hyperparameters for model training and saving checkpoints. GWAS was carried out on predictions generated by the model on a separate dataset of White British subjects consisting of 37,376 T1 images and 36,231 T2 images not included in deep learning training. Those were further divided into discovery (22,962 for both T1/T2) and replication group (12,848 T1 and 11,717 T2) for performing GWAS. See Supplementary Table 1 for sample size description.

### Deep learning architecture

Deep 3D convolutional autoencoder was used to obtain the 128-dimensional endophenotype (See Supplementary Figure 30). A separate model was trained for T1 and T2. The architecture was implemented using PyTorch and trained with the PyTorch Lightning framework. To obtain representation of the whole brain, we take the full-resolution brain T1 and T2 MRI as input. The model consisted of an initial convolutional block, four encoder blocks, a linear latent space of 128-dimension, four decoder blocks, and a final convolutional block and has 138.12 million parameters. The initial convolutional block consisted of two blocks of a 3D convolution layer (Conv3d) of kernel size 3 and stride 1 followed by a 3D batch normalization layer (BatchNorm3d) and Leaky rectified linear unit function (LeakyReLU). Each encoder block comprises a 3D max pooling layer with kernel size of 2 followed by two blocks of Conv3d of kernel size 3, BatchNorm3d, and LeakyReLU. The linear layer converts the last encoder ‘s output into 128-dimension latent space without any spatial resolution. This bottleneck vector is the representation that we are interested in as the ENDOs. Afterwards, we use the same number of decoder blocks with deconvolutional layers that gradually increase the image resolution while reducing the number of channels. Each decoder block comprises two blocks of Conv3d with kernel size 3, BatchNorm3d, and LeakyReLU, followed by 3D transposed convolution operator with stride of 2 and kernel size of 2. The final convolutional block comprises two blocks of Conv3d with kernel size 3 and stride 1, BatchNorm3d, and LeakyReLU, followed by a 3D convolutional layer with kernel size 1. The output image is of the same size as the input MRI (182×218×182).

We make the following remarks on our design choices. First, we chose a standard convolutional encoder and decoder architecture as such designs are known to deliver good performance. Second, while our architecture has semblance of the well-known U-net^24^, we do not introduce the skip connections between the encoder blocks and decoder blocks as we hope to retain maximal information through the bottleneck layer. Since we are not aiming to generate sharp images at the high resolution, the skip connections are not necessary. Third, we use the full image as input and each of the bottleneck neurons has the receptive field of the entire image. This design will ensure each endophenotype can be a descriptor of any feature across the entire brain. Fourth, we use 128 dimensional vectors as ENDOs as they are providing a comprehensive description of the entire brain morphology while making GWAS computationally feasible. But a larger number of dimensions are possible.

Compared to most previous deep learning-based brain MRI studies, the scale of our work is large both in terms of image size and sample size. Although no previous studies used an unsupervised approach for obtaining endophenotypes for GWAS for brain, some studies used autoencoder-like architecture to derive features for disease predictions such as Alzheimer’s disease^51^, schizophrenia^52^, suicidal ideation prediction^53^, and Autism spectrum disorder^54^. However, due to computational constraints, common approaches either downsize the image by reducing resolution, leading to loss of detail information, or feed the images in patches, losing the panoramic view of the complete MRI that better encodes anatomical relationship, or filter the image by extracting only gray matter and thus losing the rest of the brain information. We used a linearly registered whole-brain MRI without splitting the brain into patches to derive endophenotypes and thus offer more complete encoding of the input 3D image. Leveraging on the large sample size of UKBB brain imaging study, we have sufficient sample sizes for training a model, as well as have sufficient sample sizes for GWAS.

### Deep learning training

The dataset was randomly divided into a training set of size 4,597 images (75%) and a validation set of size 1,533 images (25%). The validation set was used for tuning hyperparameters for model training and saving checkpoints. The checkpoint with the lowest loss in the validation set was used to generate model endophenotypes.

No activation function used at the output, making it a regression task per voxel. As in the standard autoencoder, we use a reconstruction loss that will minimize the difference between the input and output images. Mean square error was used as the loss function. A mask was derived based on the input image where the background was excluded. The mean square error calculation included only the voxels corresponding to the brain region. Specifically, the loss was defined as ∑_*ijk*_(*R*(*i, j, k*) − *O*(*i, j, k*))^2^ *f*(*O*(*i, j, k*)), where *R* is the reconstructed imaging data, *O* is the original imaging data, *f* is the step function outlining the brain mask and *i, j, k* are the spatial indices. Adam optimizer was used with effectively tuned learning rate (Supplementary Note 2). Checkpoints were saved at the top 5 lowest validation loss. Both the models for T1 and T2 were trained for 75 epochs. Seven NVIDIA A100-SXM-80GB GPU cards were used for training. Each training and validation epoch took around 6 minutes.

### Characterization of endophenotypes

Our deep learning derived ENDOs are independent and can represent multiple regions of the brain. To better understand ENDOs, we used UKBB computed volume related imaging derived phenotypes (IDPs) using standard imaging software suites such as FreeSurfer and FSL. We use IDPs from the following T1 categories: Regional gray matter volumes (FAST), Subcortical volumes (FIRST), T1 structural brain. We also use T2 field ID 25781 Total volume of white matter hyperintensities (from T1 and T2_FLAIR images). To better understand T2 ENDOs, we used the volumetric estimates calculated by UKBB from T1 as both modalities from the same visit have been registered by UKBB. All IDPs are normalized by head size. The analysis performed using UKBB IDPs include UMAP (Uniform Manifold Approximation for dimension reduction)^31^, principal component analysis (PCA), t-distributed stochastic neighbor embedding (t-SNE), linear regression and logistic regression. Unsupervised dimension reduction techniques such as UMAP, PCA, t-SNE were used with default parameters. We used scikit-learn (v 0.24.2) for PCA and t-SNE and the UMAP python package (v 0.5.2) for UMAP. UKBB IDPs were used to color the 2D scatter plot of 2 components of each of the above approaches. For a continuous precomputed feature, we used rank based on percentile to make visualization possible. Sex is the only categorical feature. To better understand the deep learning derived endophenotypes, we used linear regression to predict IDPs from ENDOs. We use 10-fold cross-validation keeping only one visit for each patient to avoid data leakage. We use the mean coefficient of determination R^2^ from the 10-folds as the metric. We also use mean absolute error (MAE) for age prediction from ENDOs to make it comparable with existing literature. We used logistic regression to predict sex from ENDOs and use area under the curve (AUC) as the metric to make it comparable with existing literature.

### Decoder interpretation

256 ENDOs learn representation all over the brain. To identify regions of the brain represented by a specific ENDO, we develop Perturbation-based Decoder Interpretation (PerDI). The regions of the brain of that specific ENDO can then be associated with the SNPs identified by the same ENDO. We add one standard deviation (σ) as noise to the specific ENDO we are trying to interpret while keeping other ENDOs constant. The original decoder is used to reconstruct images from the perturbed ENDOs (perturbed reconstructed images). The process is repeated for 500 MRIs from 500 randomly selected individuals for improving the robustness of the result. Paired t-test is carried out between the 500 original reconstructed images and 500 perturbed reconstructed images. Absolute t-map is obtained. Gaussian filter (σ=3) is used to smoothen the final t-map. Using Gaussian blur reduces the impact of not using non-linear registration.

### t-map annotation

Harvard-Oxford cortical and subcortical atlas^55^ are selected to annotate the t-map. For each specific atlas, each voxel in the t-map is ranked from highest to lowest rank. For each region, normalized Kolmogorov–Smirnov test (K-S test) statistic is obtained. Specifically, the curve in Figure 3d is defined as *k*/*V* − *n*/*N*, where *k* is the number of voxels belong to a specific region in the top *n* ranked voxels, *r* is the ratio between the volume of the region and the whole brain, *V* is the number of voxels in that specific region and *N* is the total number of voxels. The higher the value of the K-S statistic, the higher the representation of the region by the specific ENDO. The brain regions corresponding to the ENDO can then be associated with the SNPs identified by the same ENDO through GWAS.

### Genome-wide association study (GWAS)

The genome-wide scans were performed over 658,720 directly genotyped SNPs (Applied Biosystems UK BiLEVE Axiom Array (field: 22438)). We used BOLT-LMM_INF (Version 2.3.4)^56^ (https://alkesgroup.broadinstitute.org/BOLT-LMM/BOLT-LMM_manual.html) for running GWAS using 256 dimension embedding obtained from T1(128 dimension) and T2(128 dimension) MRI using linear mixed model association methods. Age, gender and the first 10 ancestral principal components were used as covariates. To control for potential confounding due to ethnicity, we only include White British participants (UKBB field ID 21000: Ethnic background and field ID 22006 (Genetic ethnic grouping)) for GWAS. White British participants (35,810 T1/ 34,679 T2) not included in deep learning training are splitted into discovery (22962 T1/T2) and replication cohorts (T1=12,848/T2=11,717). The discovery cohort comprised 22,962 T1 and 22,962 T2. The replication cohort comprised 12,848 T1 and 11,717 T2.

Separate analyses were performed for both T1 and T2. Since the two modalities are related to each other, we only select the hit from the modality with more significant p-value rather than meta-analyzing the results. No genetic information was used while training the deep learning models which resulted in well controlled genomic inflation factor (Supplementary Figure 17).

We used the LDSC v 1.0.1(https://github.com/bulik/ldsc)^57^ to calculate the heritability of ENDOs. The genomic inflation factor was calculated by dividing the median chi-square statistics by the inverse cumulative distribution function of chi-square distribution of 1 degree of freedom at 0.5.

### Querying GWAS Catalog

For each locus, we used PyLiftOver 0.4 (https://github.com/konstantint/pyliftover) to transform the range from the first to the last significant SNP. UKBB uses GRCh37 as the reference genome, whereas GWAS catalog uses GRCh38. PyLiftOver converts genome coordinates between different assemblies using the UCSC liftover tool (https://genome.ucsc.edu/cgi-bin/hgLiftOver). The GWAS Catalog database (https://www.ebi.ac.uk/gwas/home) was queried using the range with additional 250 kb flanking regions. This is consistent with the approach used to create loci. We queried the GWAS catalog on November 16, 2022. Since there is no standard classification for the traits, we manually classified them into brain morphology traits, neurological disease traits, psychiatric disease traits and Alzheimer’s disease related traits based on clinical knowledge (see Supplementary Table 8 for the classification). The variants associated with Big40’s IDPs were downloaded (https://open.win.ox.ac.uk/ukbiobank/big40/), the variants were first clustered using the same 250kb threshold and then interval trees were built for each chromosome treating each cluster ± 250kb as an interval for future queries.

### Querying Big40 results

For identifying unique loci not discovered in the big40 server, we used “Table of local-peak associations (-Log10(P) > 7.5)” hosted at https://open.win.ox.ac.uk/ukbiobank/big40/. We searched for any SNP in big40 for T1/T2-FLAIR phenotypes (V0001-V1437) in the range +/- 250kb (1cM) of the loci identified by our model. We identified 13 loci which had no SNP in the 1 cM range in the big40 results.

### Meta-analysis

We used METAL (generic-metal-2011-03-25)^33^ to perform meta-analysis of GWAS summary statistics from the discovery and replication cohort.

### Genetic correlation

We used LDSC v 1.0.1 (https://github.com/bulik/ldsc)^57^ to calculate genetic correlation and LD score from the meta-analysis results. We computed genetic correlations for attention-deficit/hyperactivity disorder (ADHD), Alzheimer’s disease (AD), amyotrophic lateral sclerosis (ALS), autism, bipolar disorder (BPD), ischemic stroke, major depressive disorder (MDD), neuroticism, schizophrenia (SCZ), and sleep disorder.

## Data Availability

We share our code and model checkpoints at https://github.com/ZhiGroup/DeepENDO. All results are available at https://deependo.org.

## Data availability

We share our code and model checkpoints at https://github.com/ZhiGroup/DeepENDO. All results are available at http://deependo.org.

## Author contributions

Z.X. and D.Z. conceptualized the design of the study. K.P., Z.X., and H.Y. designed and implemented the model. K.P. and Z.X. led the design of the analysis strategy and pipeline, with substantial input from H.C., L.G., A.K., E.F., and M.F.. K.P. and Z.X. trained the model, generated predictions and characterized the endophenotypes, and conducted GWAS. K.P., Z.X. and D.Z. designed and implemented the decoder-based visualization, with substantial inputs from L.G. and E.F.. K.P., Z.X., S.M.S.I., W.Z., and A.G. conducted post GWAS analyses. K.P. Z.X. and D.Z. led the result interpretation, with substantial inputs from S.M.S.I., A.F., L.G., E.F. and M.F.. E.F., M.F., S.J., and D.Z. secured the funding of the project. D.Z. oversaw the execution of the entire project, with substantial input from M.F. and S.J.. K.P. and D.Z. led the writing of the manuscript, with substantial inputs from Z.X., S.M.S.I. and A.G.. All authors discussed, commented and confirmed the final version of the manuscript.

## Acknowledgements

This work was supported by grants from the National Institute of Aging (U01 AG070112-01A1). In addition, L.G. is supported in part by NIH grants UL1TR003167 and R01NS121154.

## Extended Data Figures

**Extended Data Figure 1.**
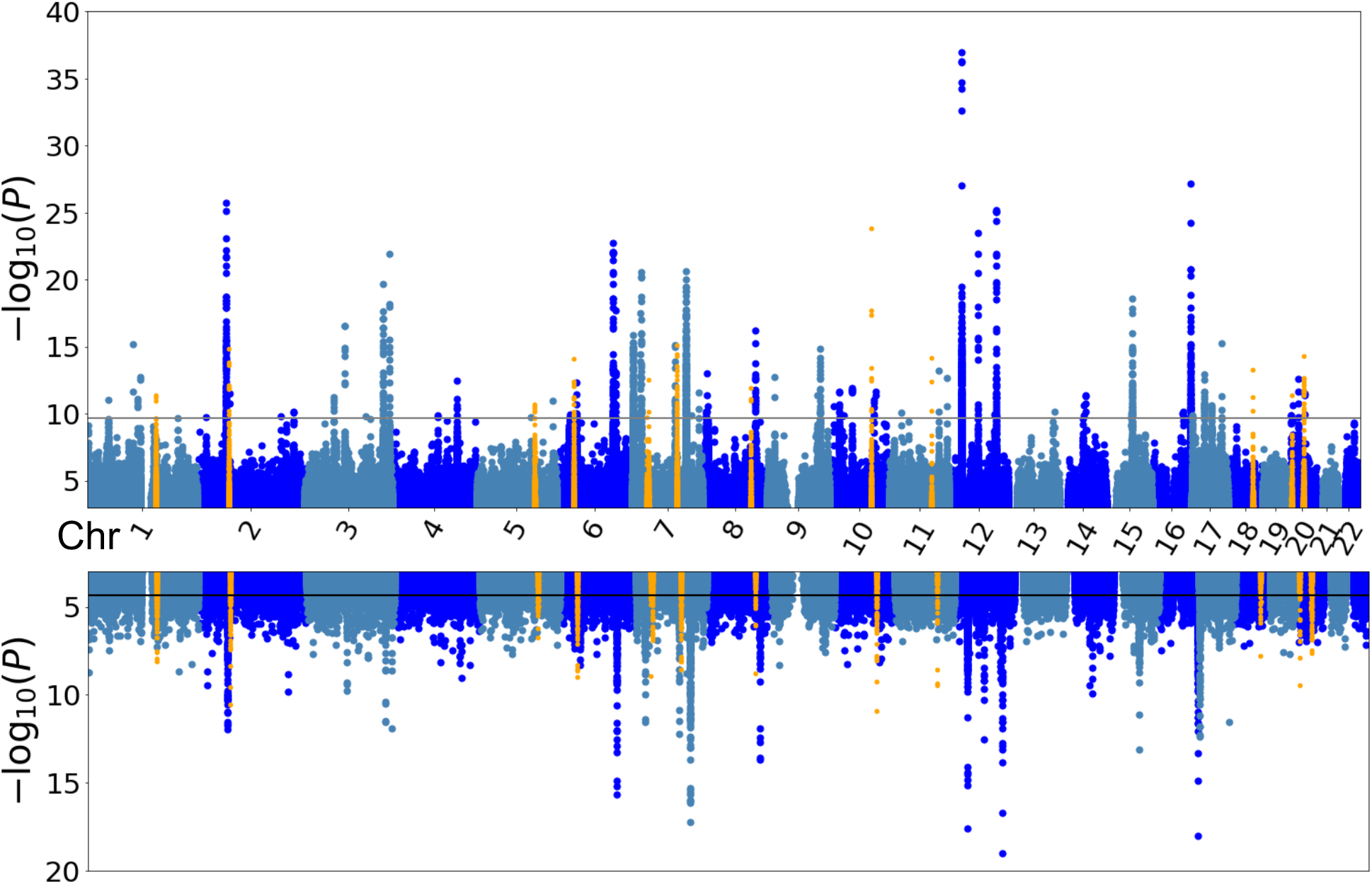
Miami plot of GWAS results in discovery and replication cohorts. Aggregated Miami plot of all 256 single ENDO GWASs is shown. Top panel is the Manhattan plot from the GWAS of the discovery cohort, the black line marks the genome-wide significance level at 5e-8/256=1.953e-10. Bottom panel is the Manhattan plot from the GWAS of the replication cohort, the black line marks the significance level at 0.05/1133 = 4.417e-5, where 1133 is the number of significant SNP-ENDO pairs.13 loci not reported in earlier UK Biobank T1 and T2 IDP GWAS are colored yellow.

**Extended Data Figure 2.**
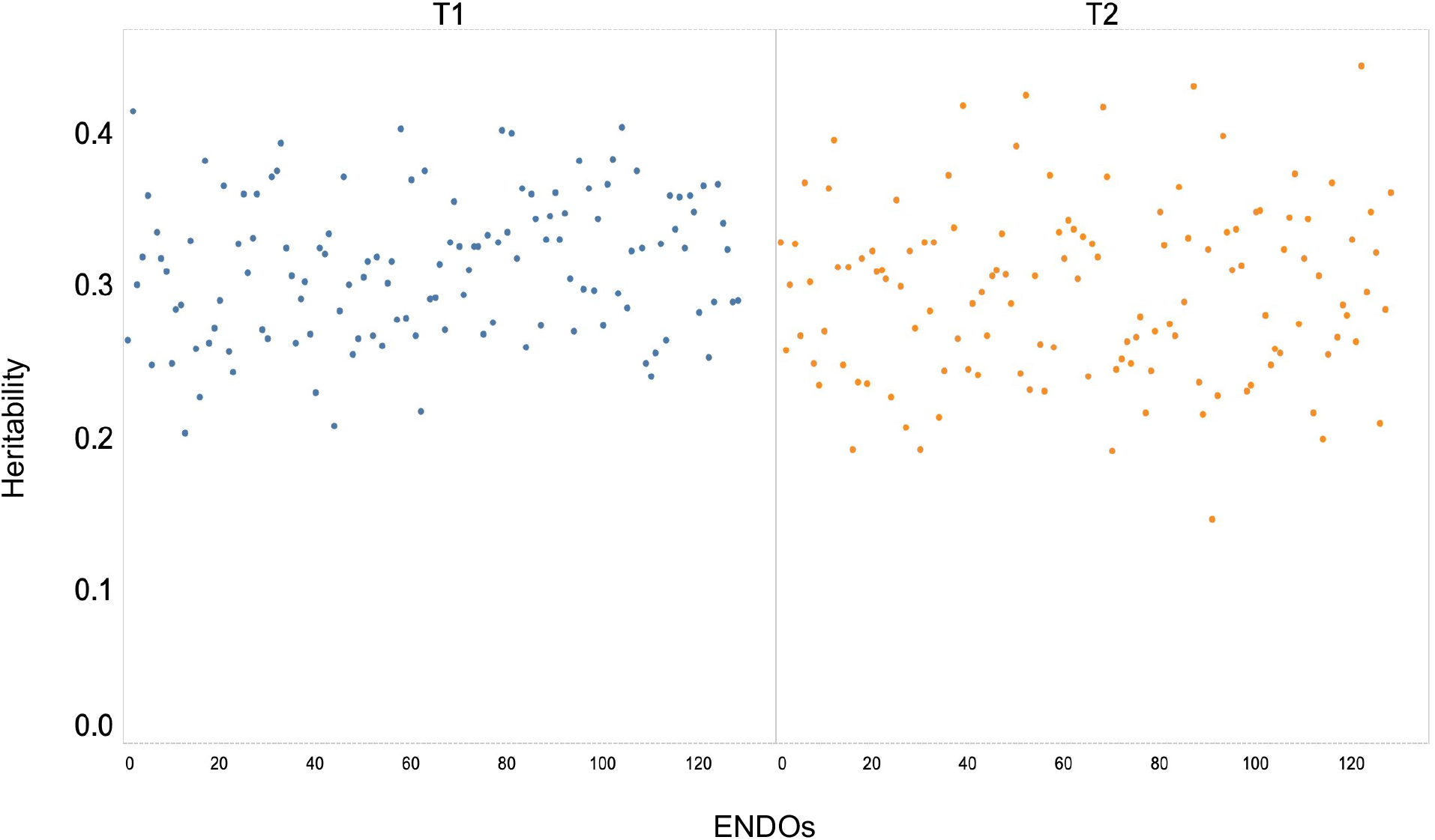
Heritabilities of ENDOs. Heritability is calculated using LDSC.

**Extended Data Figure 3.**
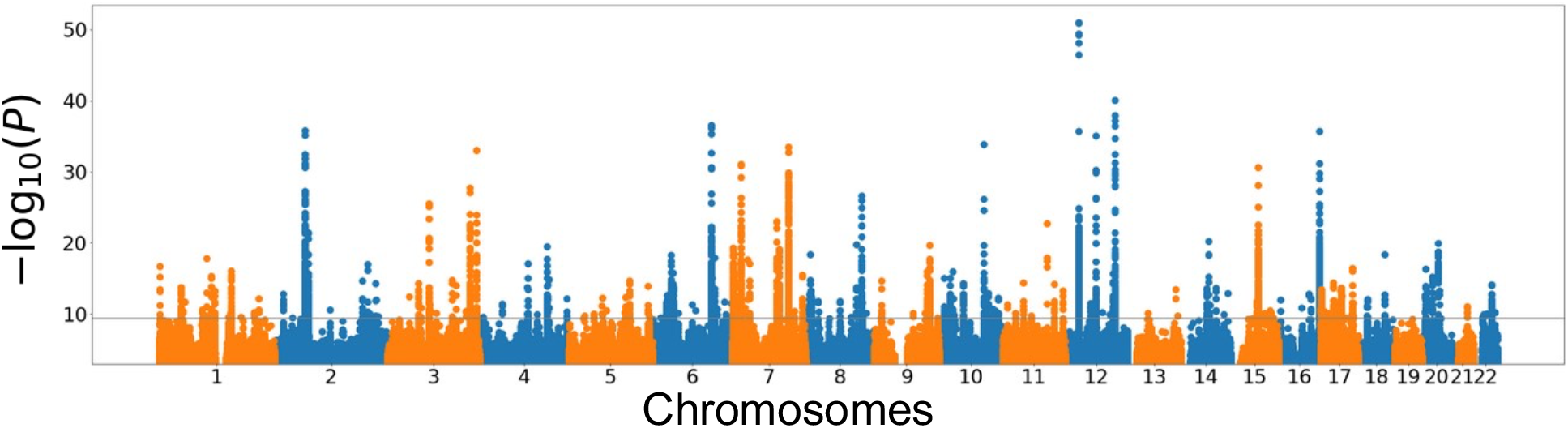
Meta-analysis Manhattan plot. Inverse-variance weighted fixed-effect meta-analysis of the discovery and the replication GWAS summary statistics discovered 3485 significant (P<5×10^−8^/256) SNP-ENDO pairs spanning. A total of 799 SNPs identified are clustered into 170 loci.

**Extended Data Figure 4.**
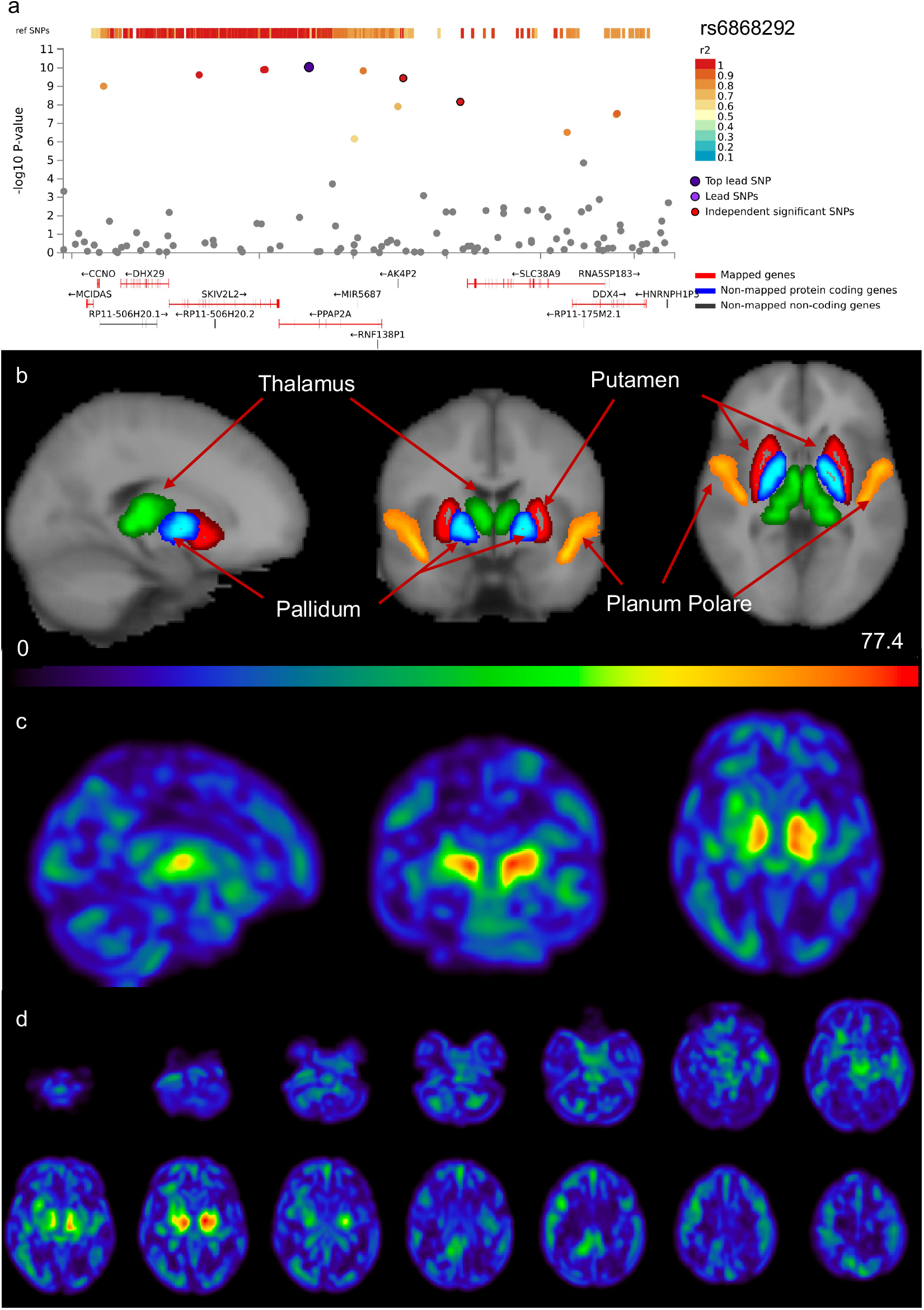
Lead ENDO T2:93 identifies a new locus on chromosome 5 that was not previously associated with brain-related traits. a) Regional plot for lead SNP rs6868292 is identified by meta-analysis. b) Harvard-Oxford structural atlas is utilized for region annotation. c) t-map shows pallidum, putamen, and thalamus as the most prominent subcortical structures, whereas planum polare is the most prominent cortical structure. d) Lightbox view (axial) displays t-map slices across the entire brain.

**Extended Data Figure 5.**
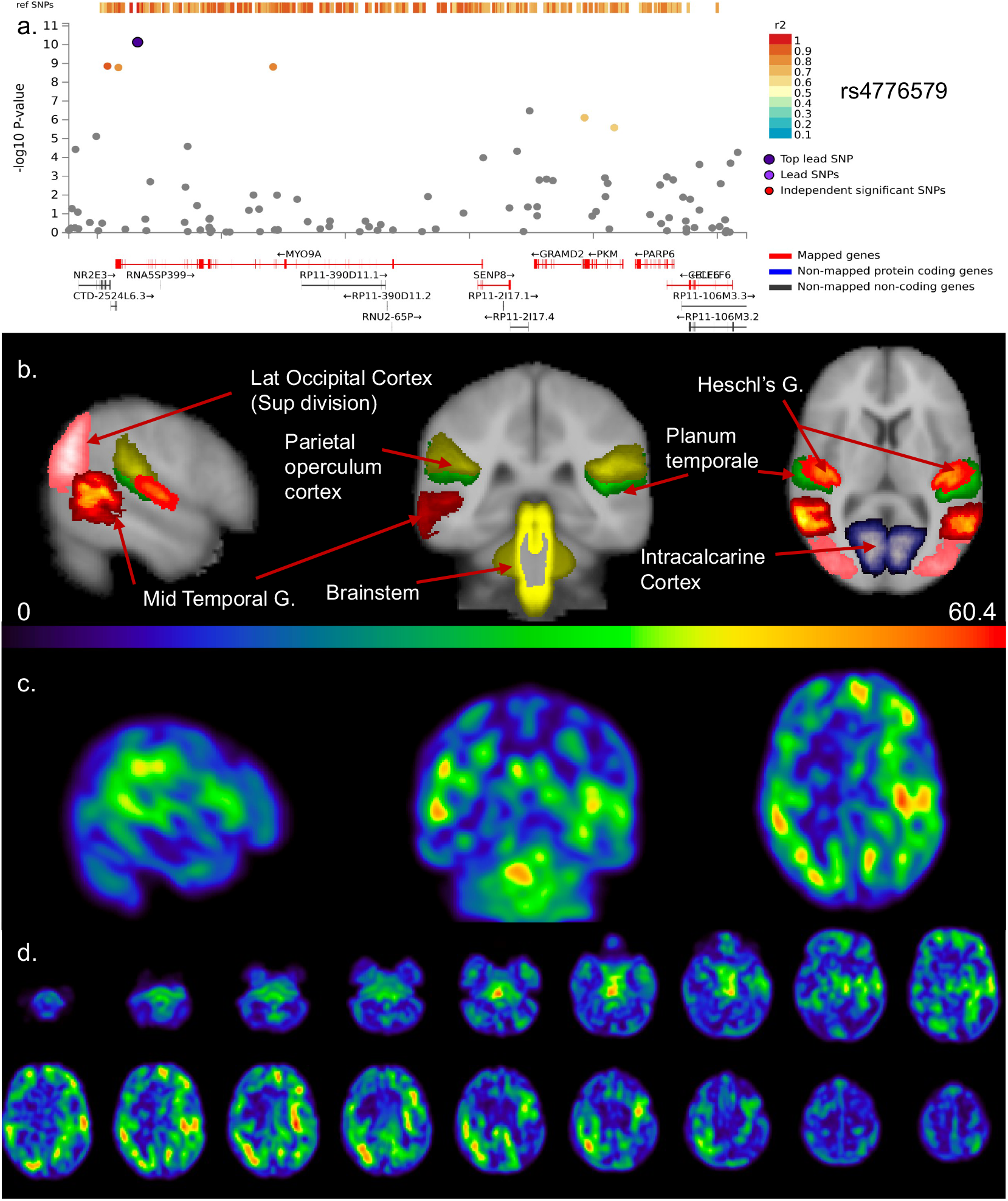
Lead ENDO T2:66 identifies a new locus on chromosome 15 that was not previously associated with brain-related traits. a) Regional plot for lead SNP rs4776579 is identified by meta-analysis. b) Harvard-Oxford structural atlas is utilized for region annotation. c) t-map shows lateral occipital cortex, middle temporal gyrus (temporo-occipital part), parietal-operculum cortex, planum temporale, Heschl’s gyrus, intra calcarine cortex, and brainstem as prominent regions. d) Lightbox view (axial) displays t-map slices across the entire brain.

