## Supplementary figures for "New phenotype discovery method by unsupervised deep representation learning empowers genetic association studies of brain imaging"

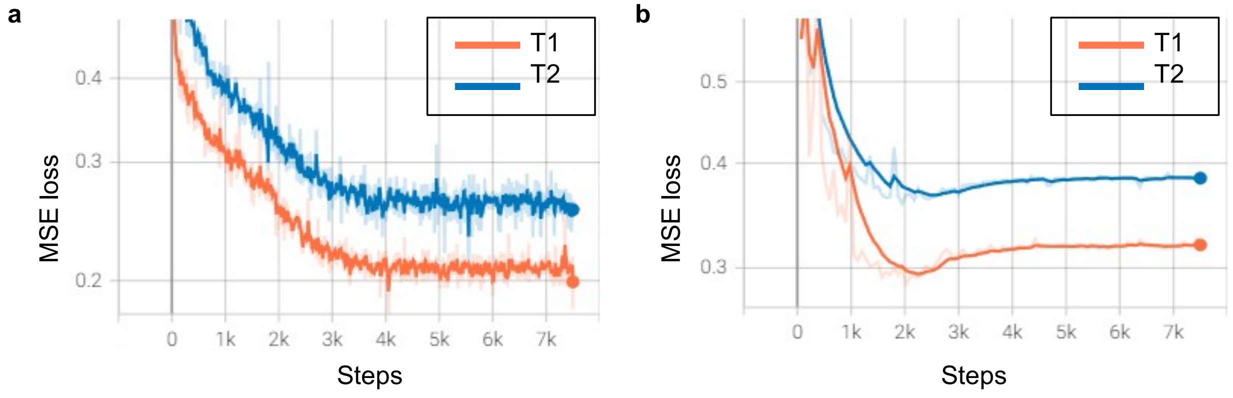


**c**

| **Modality** | **Train** (T1:n = 4,597, T2:n = 4,597) | **Validation** (T1:n =1,533, T2:n =1,533) | **Test** (T1:n=37,376, T2:n=36,231) | **Random pairs of original and reconstructed images in test set** |
|---|---|---|---|---|
| **T1** | 0.250 (0.022) | 0.291 (0.033) | 0.295 (0.032) | 0.528 (0.106) |
| **T2 FLAIR** | 0.322 (0.038) | 0.360 (0.046) | 0.362 (0.045) | 0.669 (0.124) |

**Supplementary Figure 1. Voxel-wise mean squared error (MSE) (squared L2 norm) loss focusing on the brain while masking out the background.**
Test set is not included in the deep learning training and is used for defining discovery and replication cohorts for GWAS. a) Training loss curve b) Validation loss curve. Model weights corresponding to lowest validation loss were selected c) MSE loss for train, validation and test set. MSE loss between random pairs of original and reconstructed images in the test set shows the model is learning brain morphology specific to individuals.

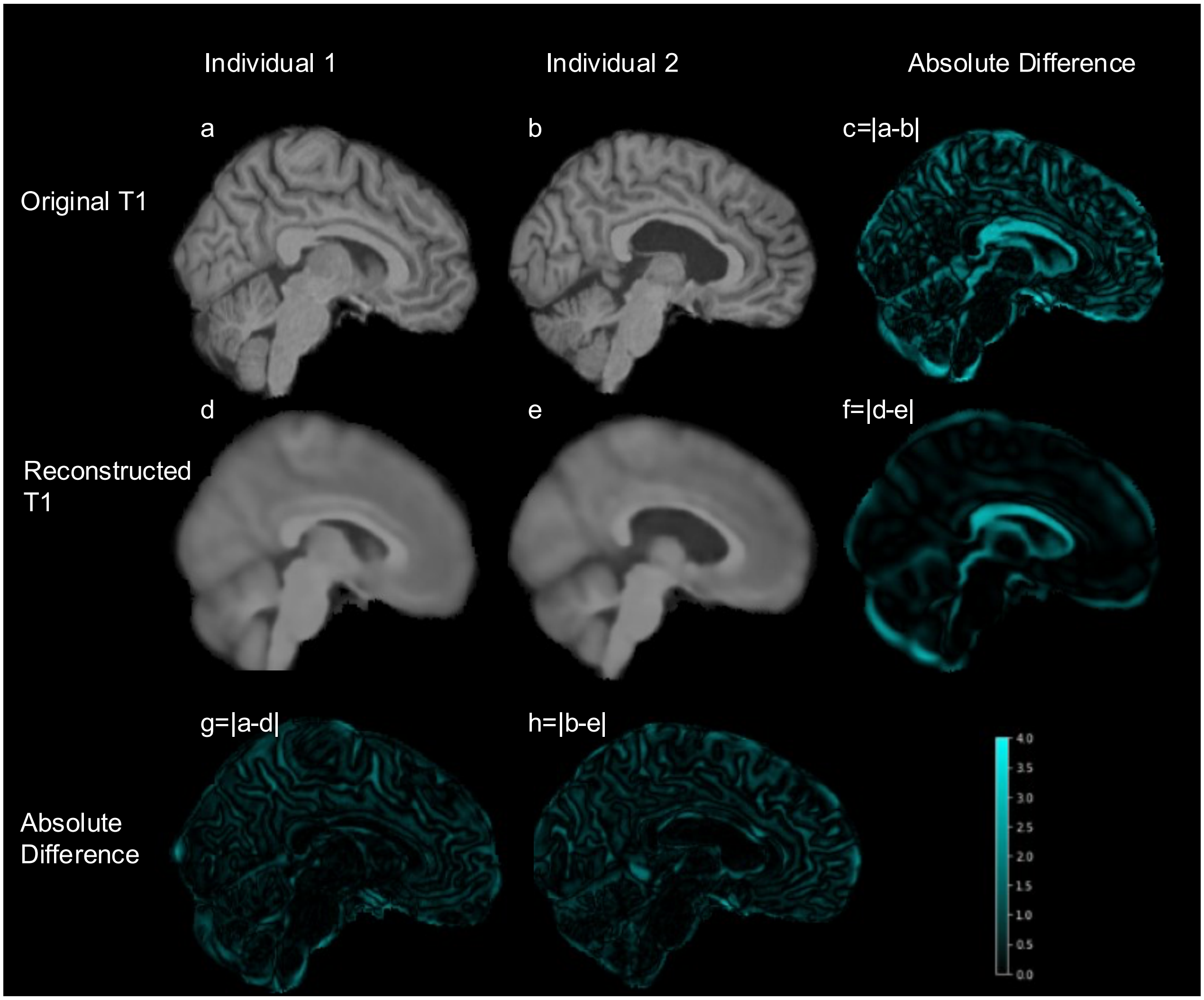


**Supplementary Figure 2. ENDOs capture variations between individuals**.
Original (a,b) and reconstructed (d,e) MRIs from two randomly selected individuals are shown. High frequency information may be lost in reconstruction due to absence of skip connections between encoder and decoder to maximize information retention in ENDOs. However, most of the overall structural details are retained. Absolute difference between two original T1 is shown (c=|a-b|) and absolute difference between two reconstructed T1 (f=|d-e|) are much greater between two individuals than between the original and the reconstructed MRIs for each individual (g=|a-d|,h=|b-e|). Model captured variance in morphology between individual subjects. Lightbox view representing multiple axial view slices in Supplementary Figure 3 (T1) and Supplementary Figure 4 (T2-FLAIR).

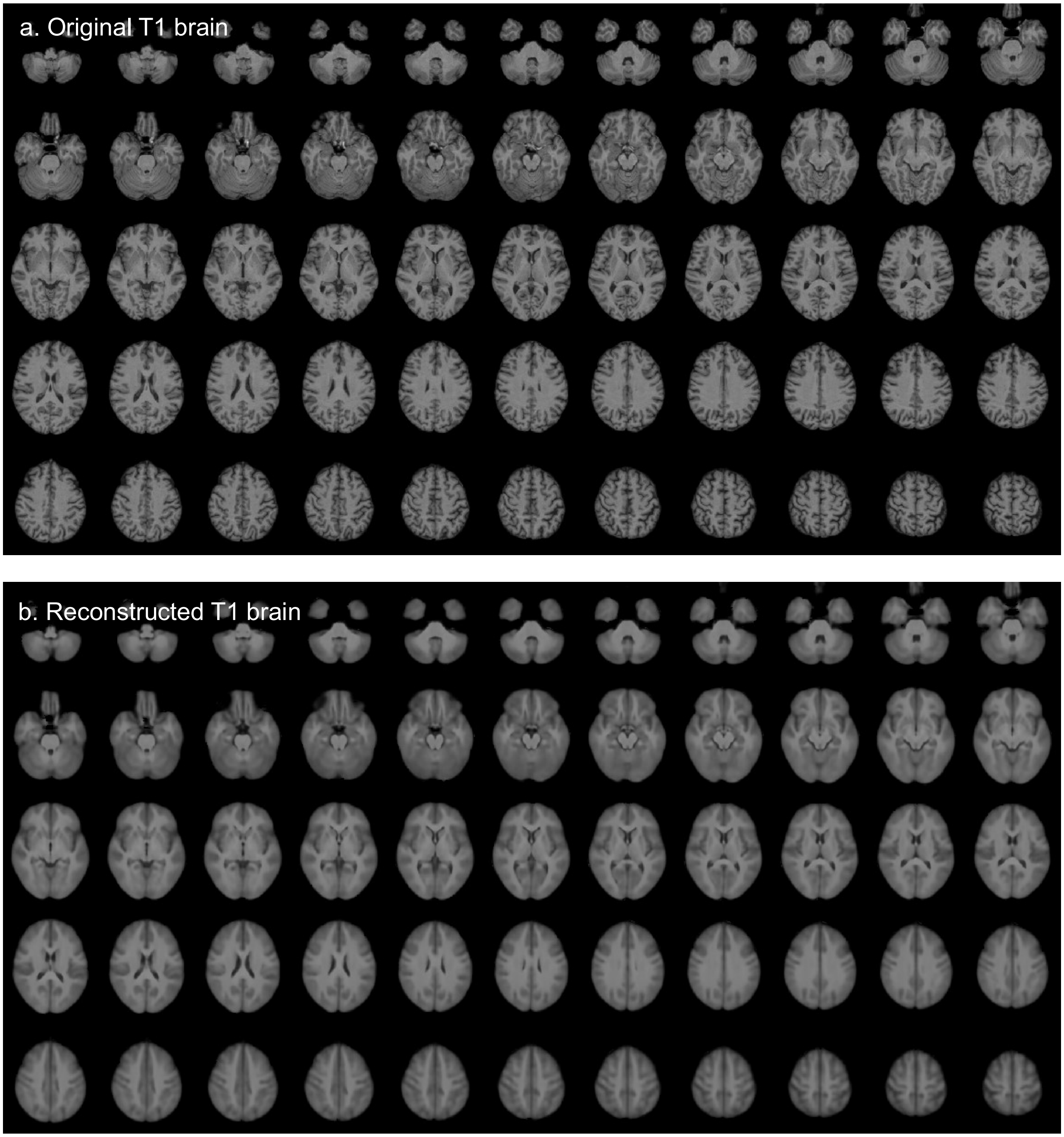


**Supplementary Figure 3. Axial view of a) Original T1 MRI b) Reconstructed T1 MRI c) Original T1 MRI d) Reconstructed T1 MRI**

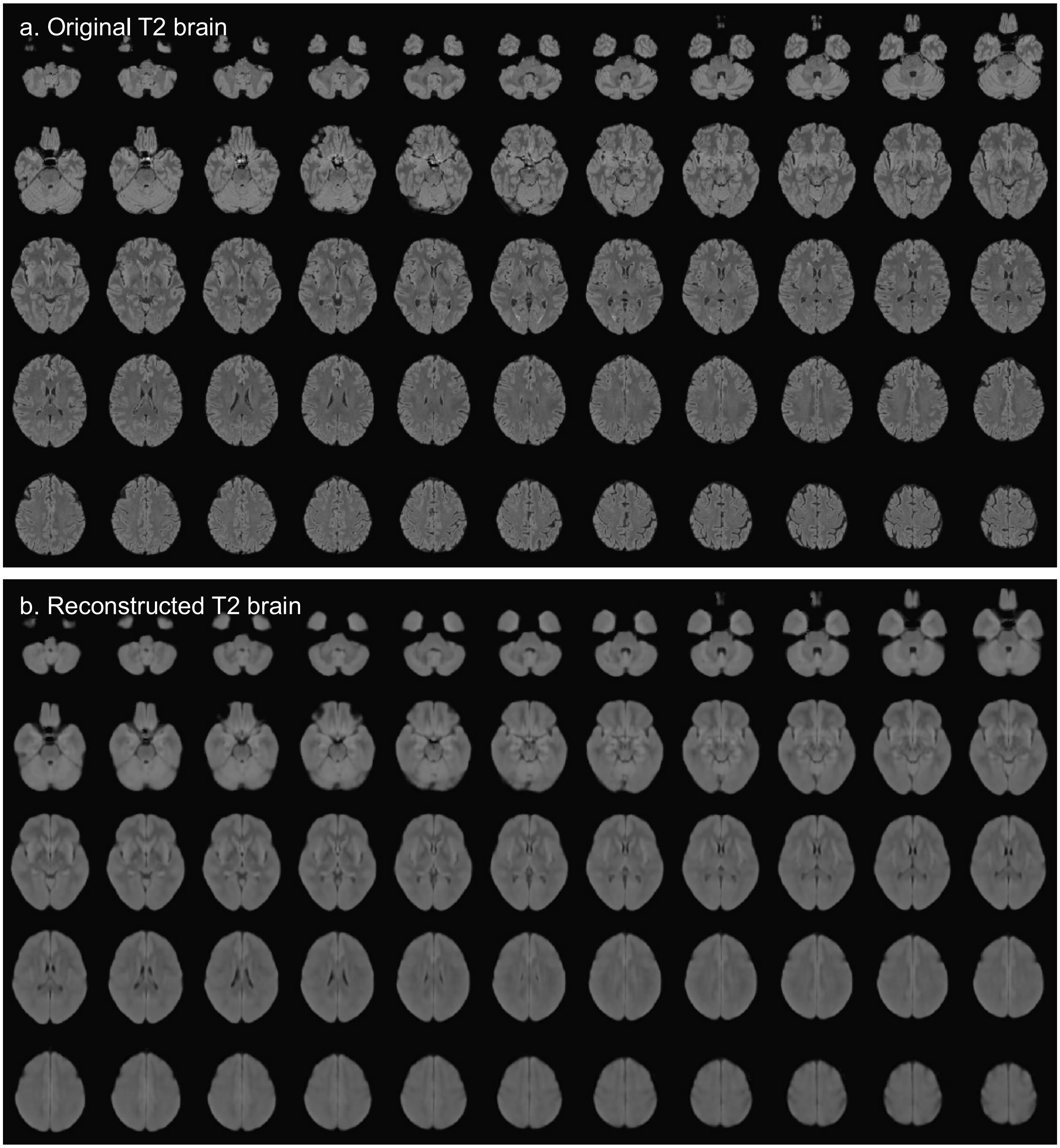


**Supplementary Figure 4. Axial view of a) Original T2-FLAIR MRI b) Reconstructed T2-FLAIR MRI**

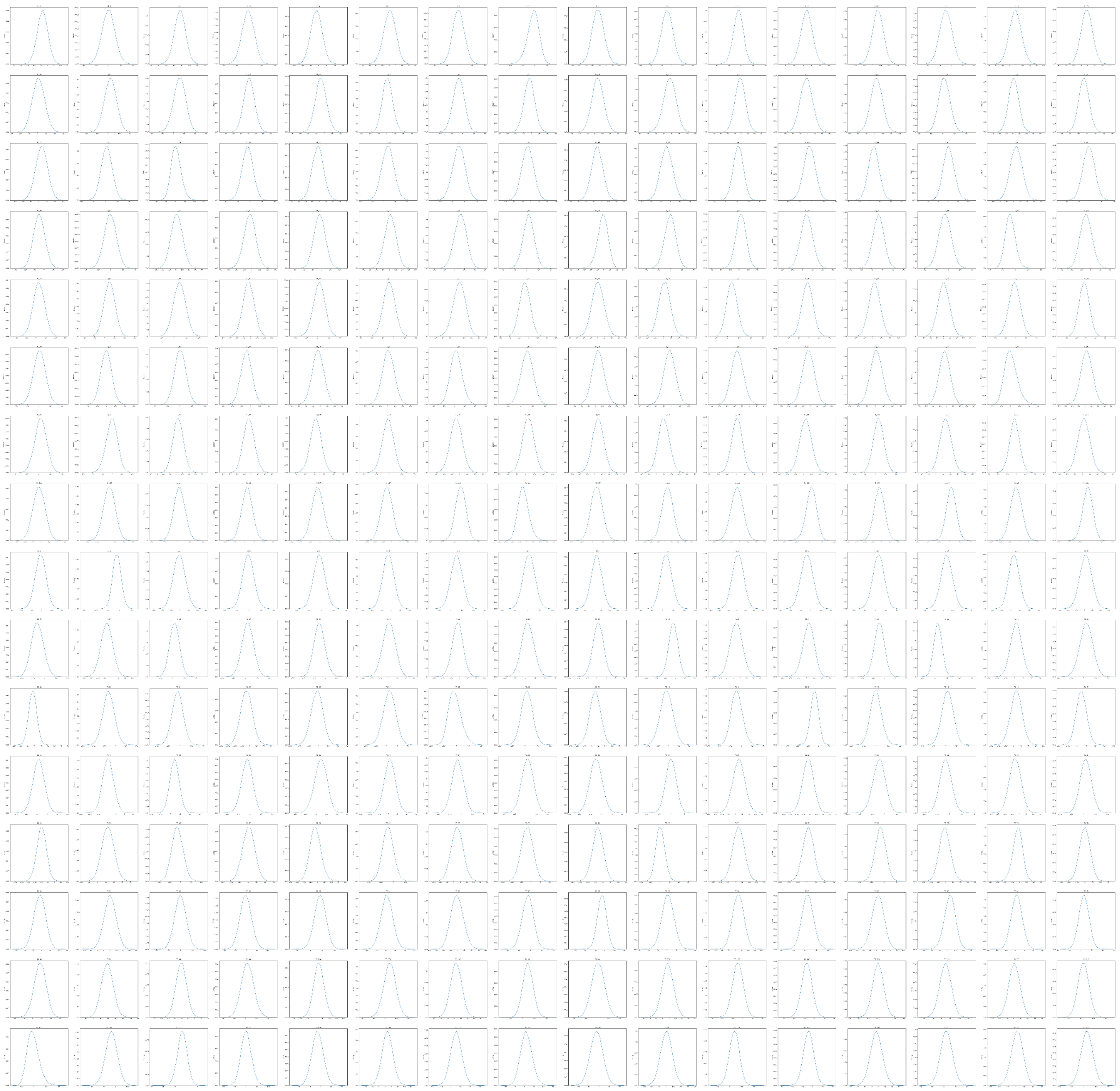

**Supplementary Figure 5. KDE plot of 256 dimensions of the ENDOs showing normal distribution of each dimension.**

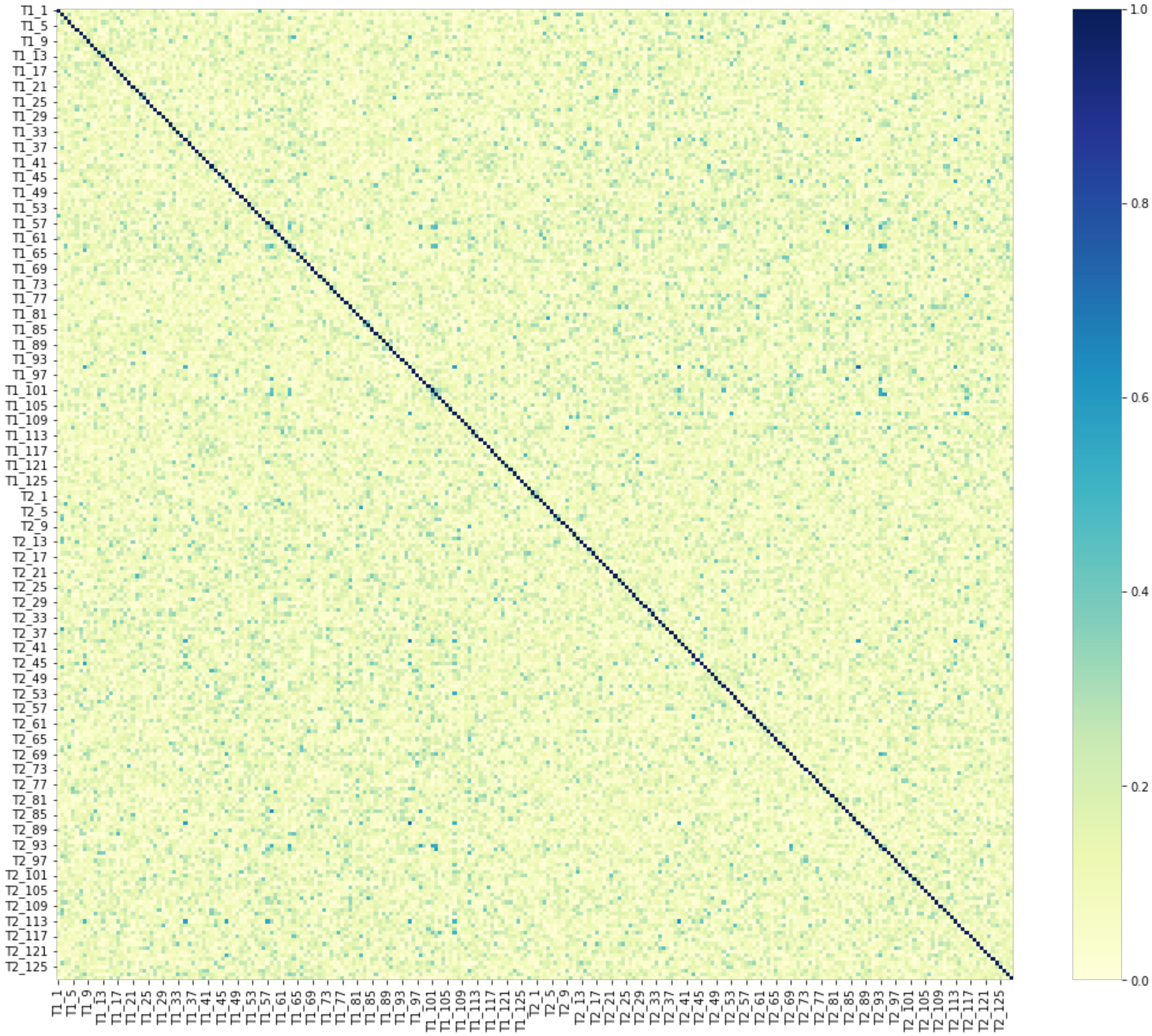


**Supplementary Figure 6. Correlation heatmap using absolute values of Pearson correlation showing independence of the ENDOs.**

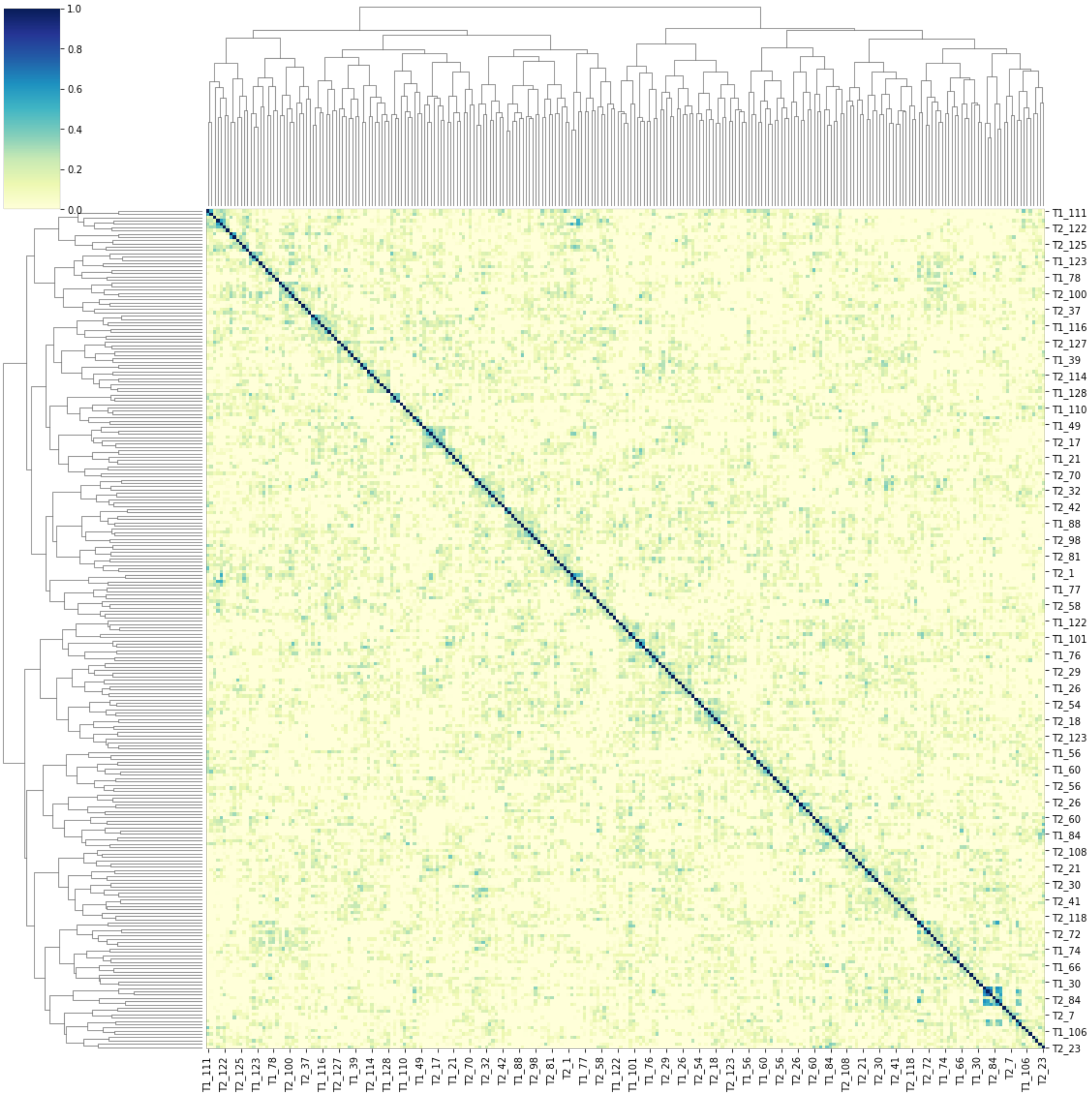


**Supplementary Figure 7. Correlation heatmap with hierarchical clustering using absolute values of Pearson correlation showing lack of clustering of the ENDOs.**

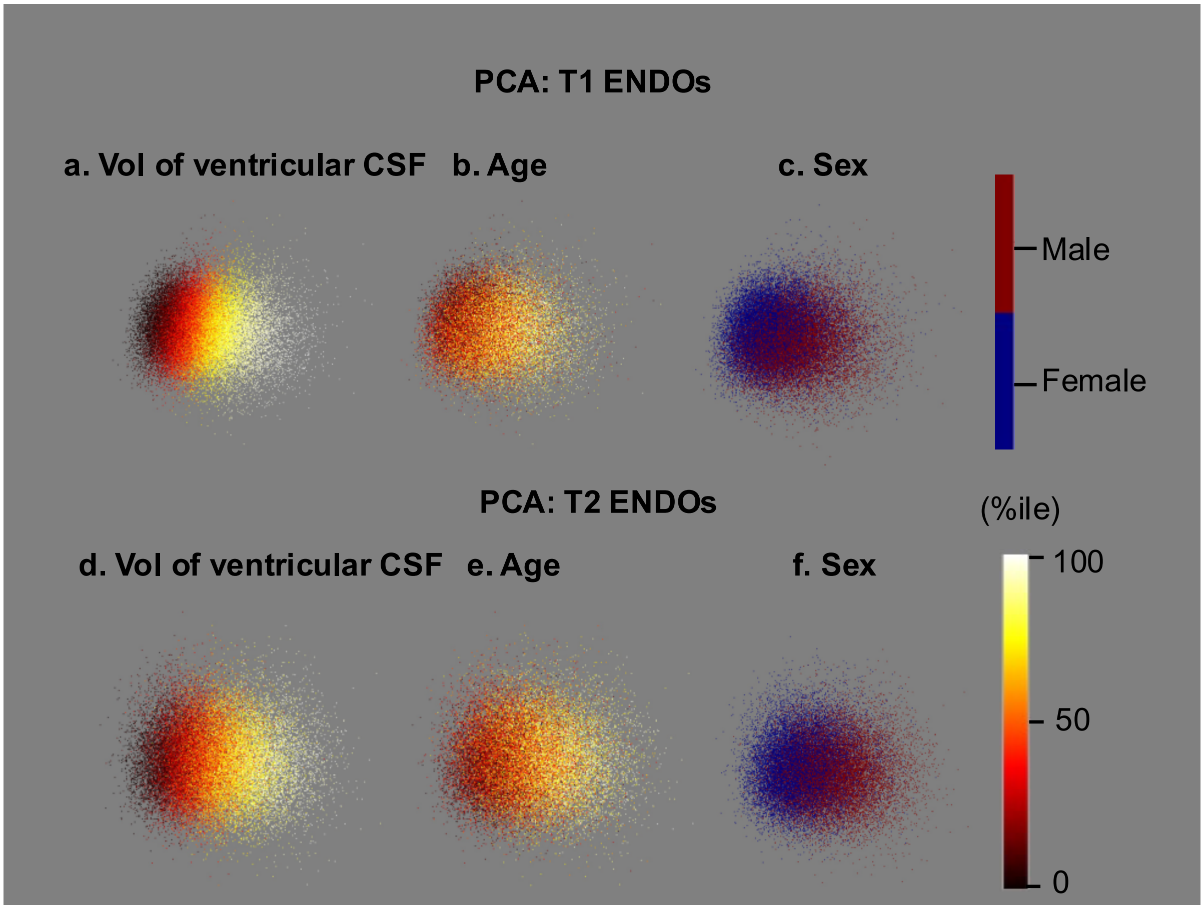


**Supplementary Figure 8. Principal component analysis (PCA) dimension reduction for ENDOs.**

ENDOs of 37,376 T1 and 36,231 T2-FLAIR MRIs of white British individuals (deep learning test set) are correlated with demographic and brain volume measures as visualized by PCA. The points are individual participants colored by UKBB provided precomputed features. For a continuous precomputed feature, we convert the value to percentile to make visualization possible. Sex is the only categorical feature.

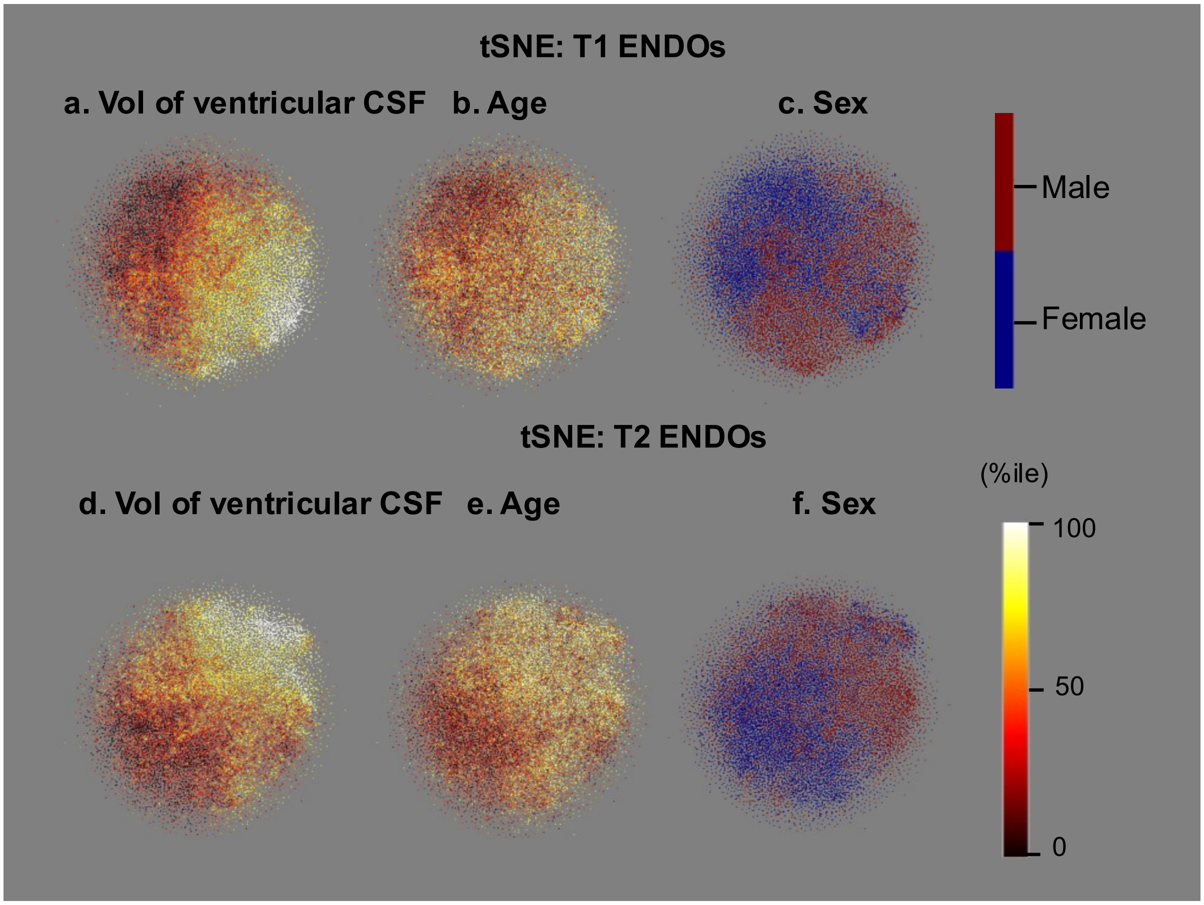


**Supplementary Figure 9. t-distributed Stochastic Neighbor Embedding (t-SNE) dimension reduction for ENDOs.**

ENDOs of 37,376 T1 and 36,231 T2-FLAIR MRIs of white British individuals (deep learning test set) are correlated with demographic and brain volume measures as visualized by t-SNE. The points are individual participants colored by UKBB provided precomputed features. For a continuous precomputed feature, we convert the value to percentile to make visualization possible. Sex is the only categorical feature.

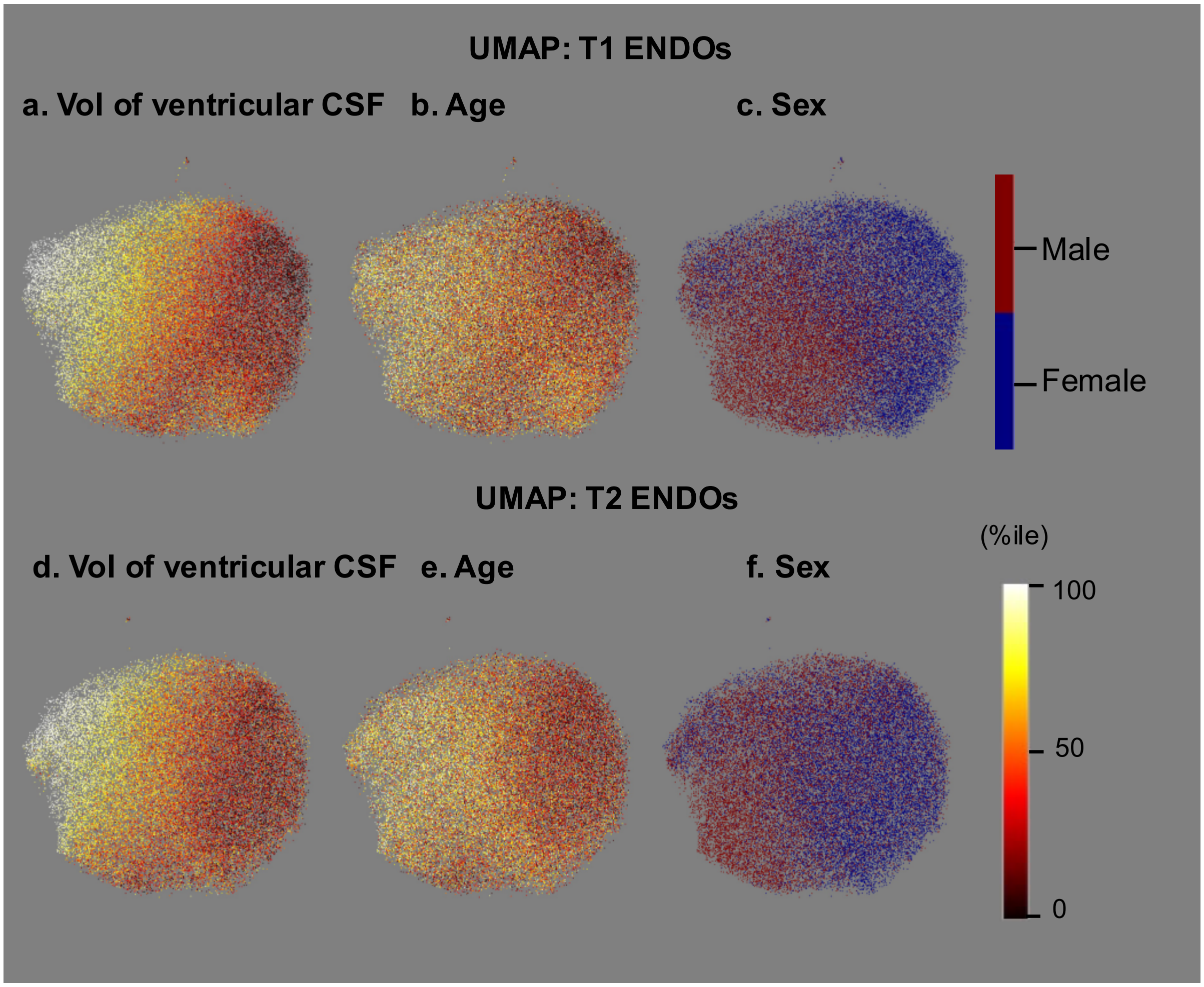


**Supplementary Figure 10. Uniform Manifold Approximation and Projection (UMAP) for dimension Reduction of ENDOs.**

ENDOs of 37,376 T1 and 36,231 T2-FLAIR MRIs of white British individuals (deep learning test set) are correlated with demographic and brain volume measures as visualized by UMAP. The points are individual participants colored by UKBB provided precomputed features. For a continuous precomputed feature, we convert the value to percentile to make visualization possible. Sex is the only categorical feature.

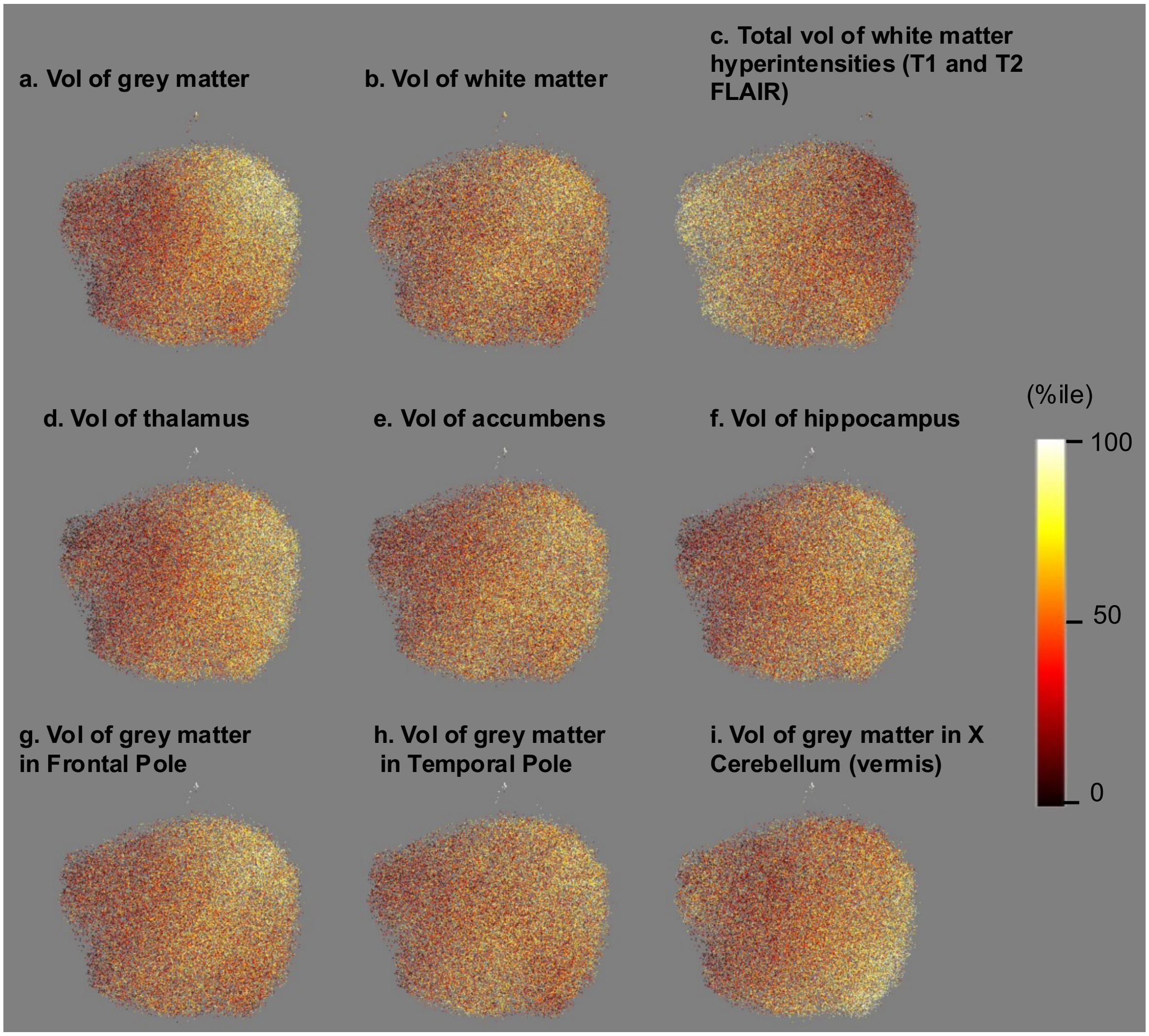


**Supplementary Figure 11. UMAP of ENDOs derived from T1.** ENDOs of 37,376 T1 MRIs of white British individuals (deep learning test set) are correlated with demographic and brain volume measures as visualized by UMAP. The points are individual participants colored by UKBB provided precomputed features. For a continuous precomputed feature, we convert the value to percentile to make visualization possible. ENDOs of 36,231 T1 MRIs of white British individuals are used for brain volume measure "total volume of white matter hyperintensities" as it is calculated from both T1 and T2. UMAP points colored by volume of ventricular CSF, age and sex are shown in Supplementary Figure 10.

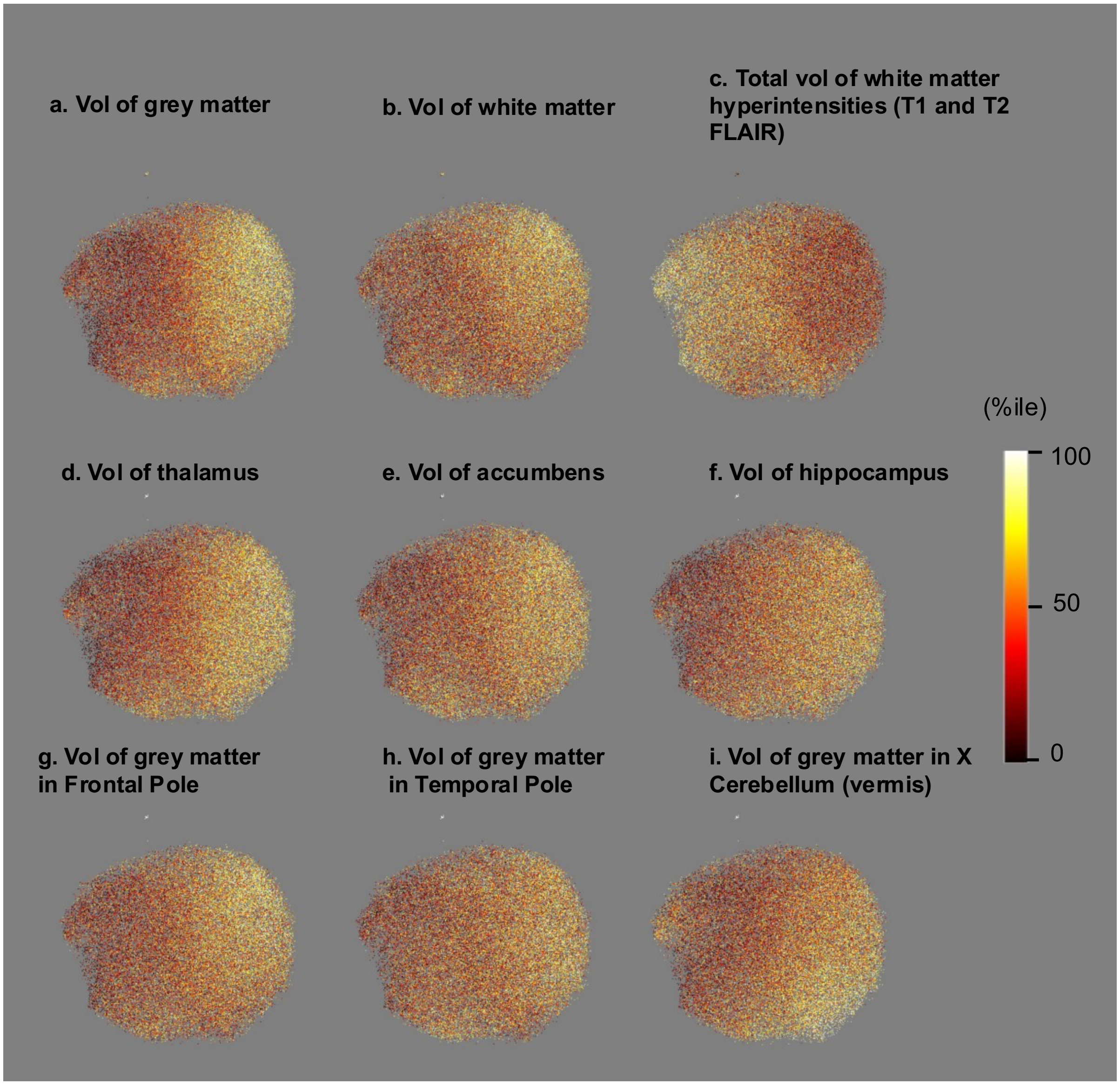


**Supplementary Figure 12. UMAP of ENDOs derived from T2-FLAIR.** ENDOs of 36,231 T2-FLAIR MRIs of white British individuals (deep learning test set) are correlated with demographic and brain volume measures as visualized by UMAP. The points are individual participants colored by UKBB provided precomputed features. For a continuous precomputed feature, we convert the value to percentile to make visualization possible. UMAP points colored by volume of ventricular CSF, age and sex are shown in Supplementary Figure 10.

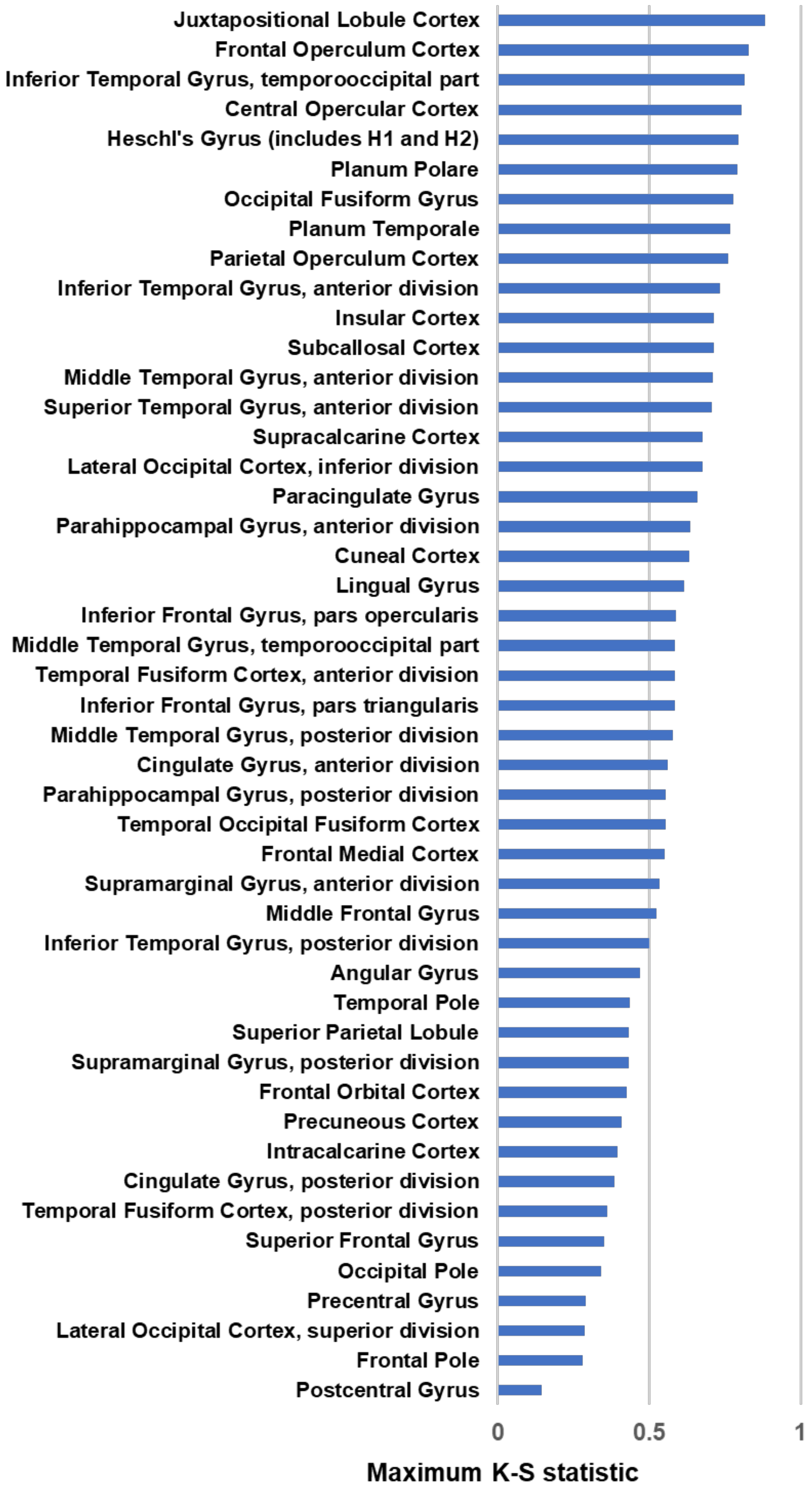


**Supplementary Figure 13. Maximum K-S statistic value across all cortical regions in T1.** Harvard Oxford cortical atlas (included in FSL) was used to rank voxels in t-map and generate K-S statistic plots.

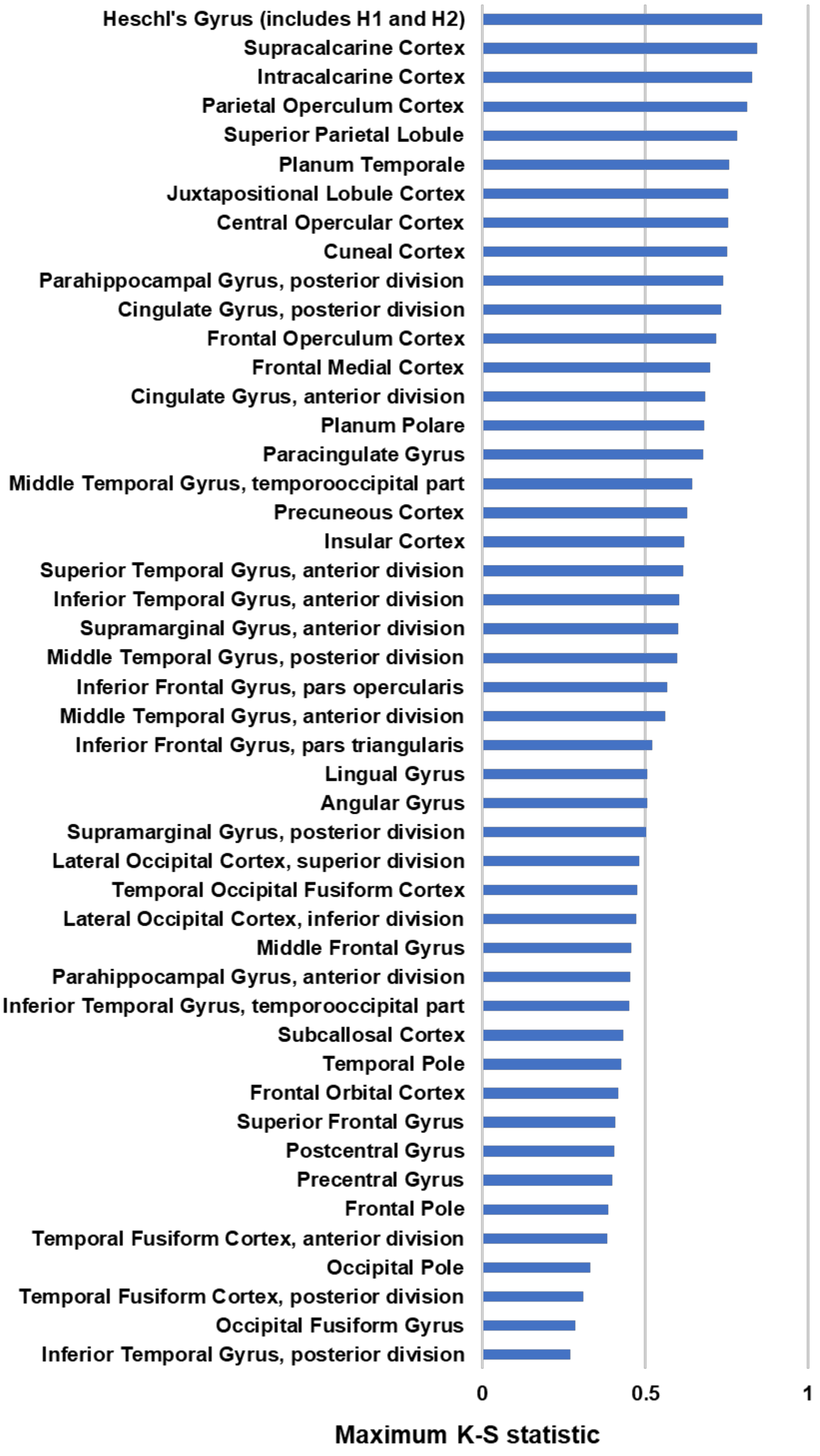


**Supplementary Figure 14. Maximum K-S statistic value across all cortical regions in T2-FLAIR.**

Harvard Oxford cortical atlas (included in FSL) was used to rank voxels in t-map and generate K-S statistic plots.

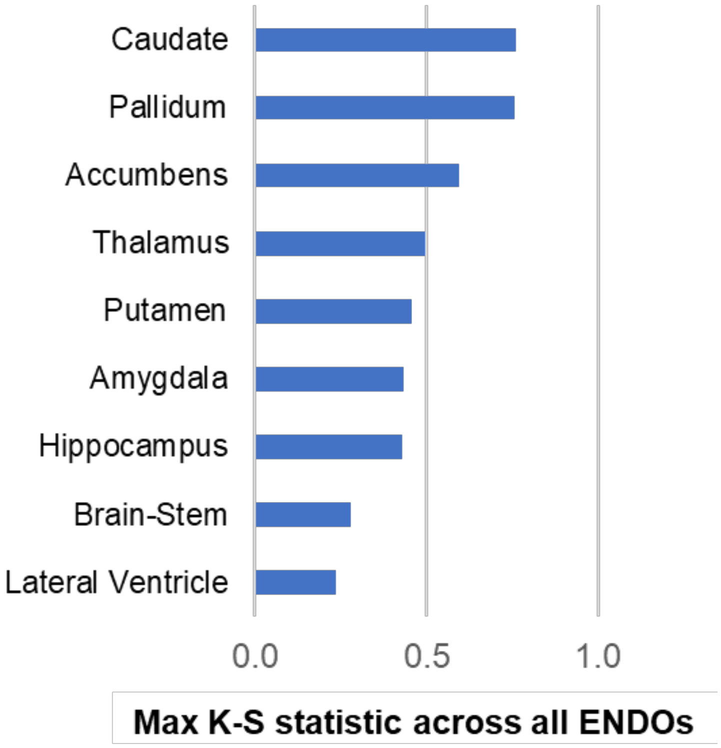


**Supplementary Figure 15. Maximum K-S statistic value across all subcortical regions in T1.**

Harvard Oxford subcortical atlas (included in FSL) was used to rank voxels in t-map and generate K-S statistic plots. Caudate, pallidum, accumbens, thalamus, amygdala, hippocampus and putamen have maximum K-S statistic value of >0.5 and are represented well.

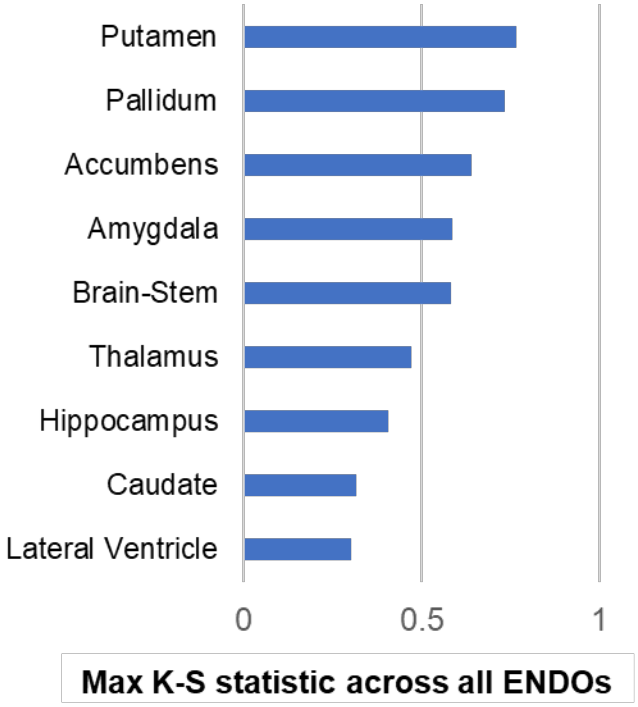


**Supplementary Figure 16. Maximum K-S statistic value across all subcortical regions in T2.**

Harvard Oxford subcortical atlas (included in FSL) was used to rank voxels in t-map and generate K-S statistic plots. Caudate, pallidum, accumbens, thalamus, amygdala, hippocampus and putamen have maximum K-S statistic value of >0.5 and are represented well.

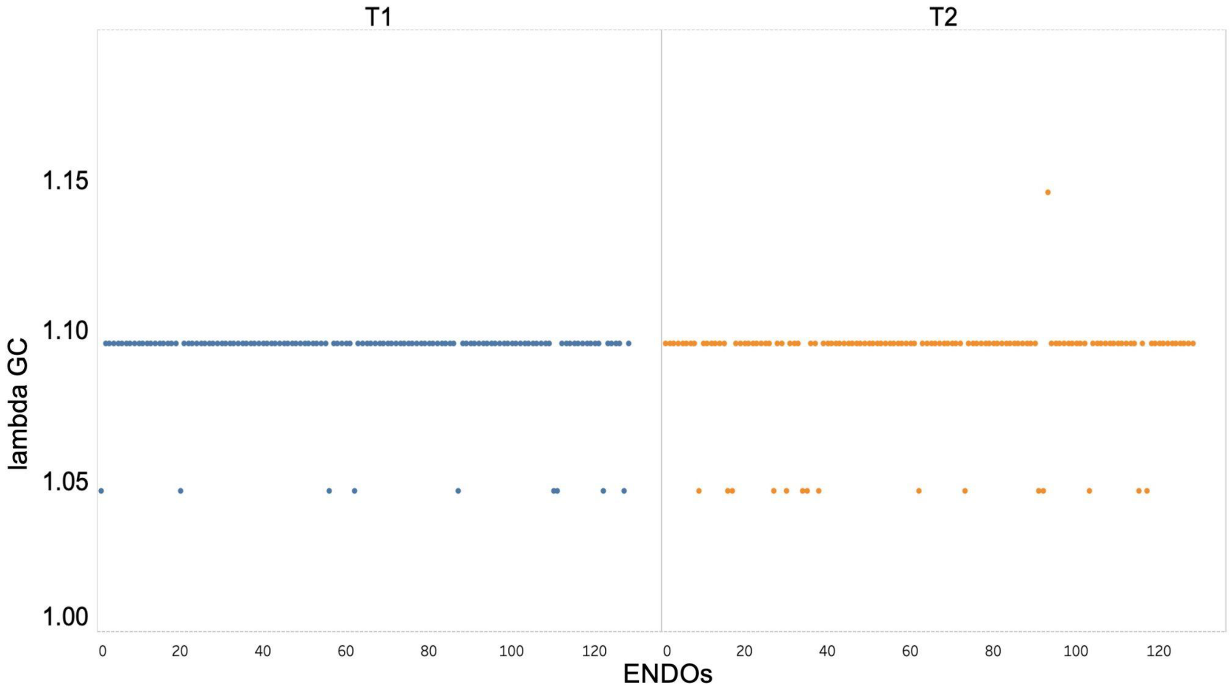


**Supplementary Figure 17. Genomic inflation factor (lambda GC).**
No genetic information was used while training the deep learning models which resulted in well controlled genomic inflation factor.

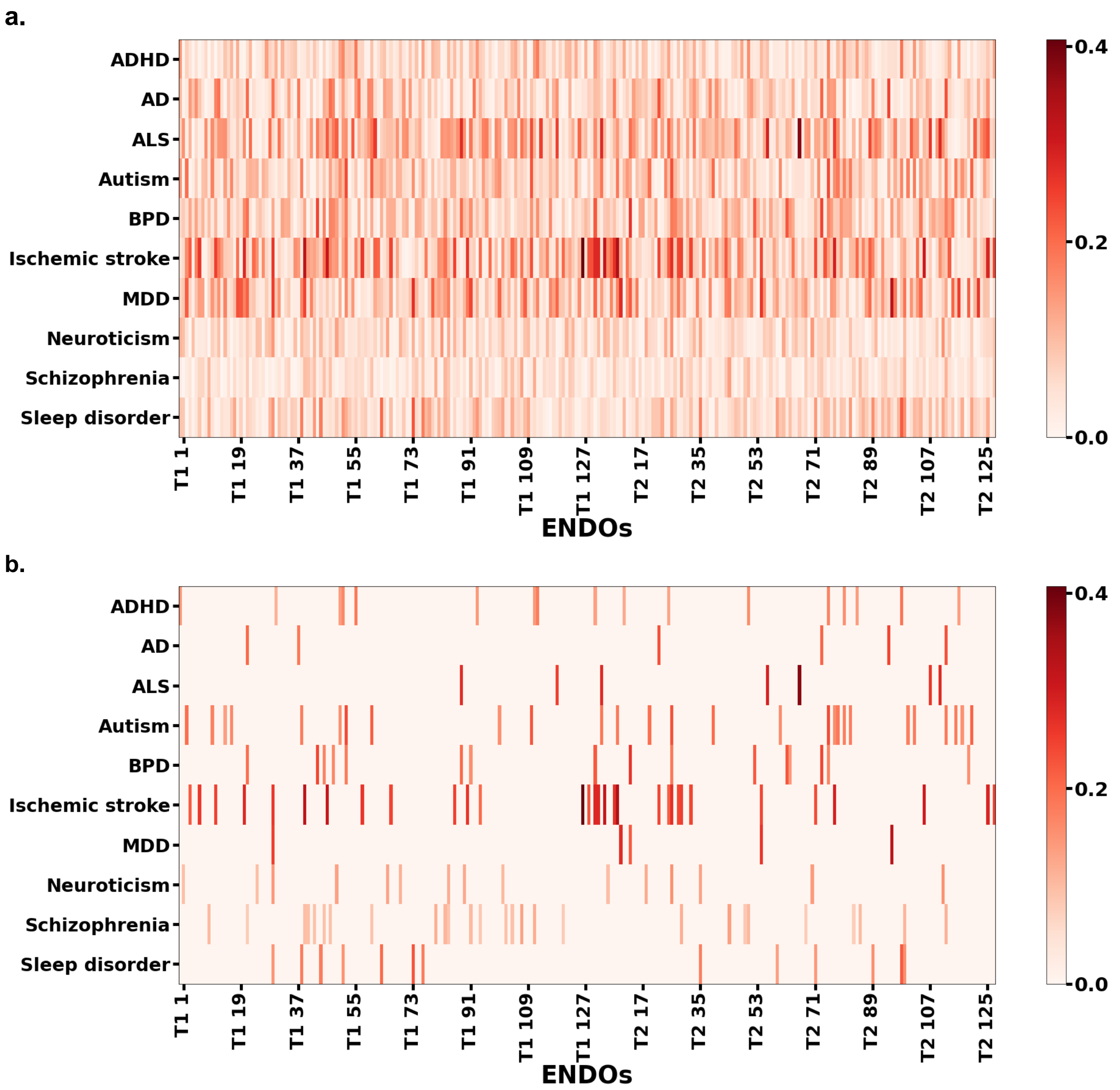


**Supplementary Figure 18. Genetic correlation of common brain-related diseases with ENDOs using meta-analysis results (Discovery and replication cohort).**

a. Mean absolute genetic correlation with various disorders using meta-analysis results. b. Mean absolute genetic correlation having p <0.05

ADHD: Attention-deficit/hyperactivity disorder, ALS: Amyotrophic lateral sclerosis, BPD: Bipolar disorder, IS: Ischemic stroke, MDD: Major depressive disorder, Sleep: Sleep disorder

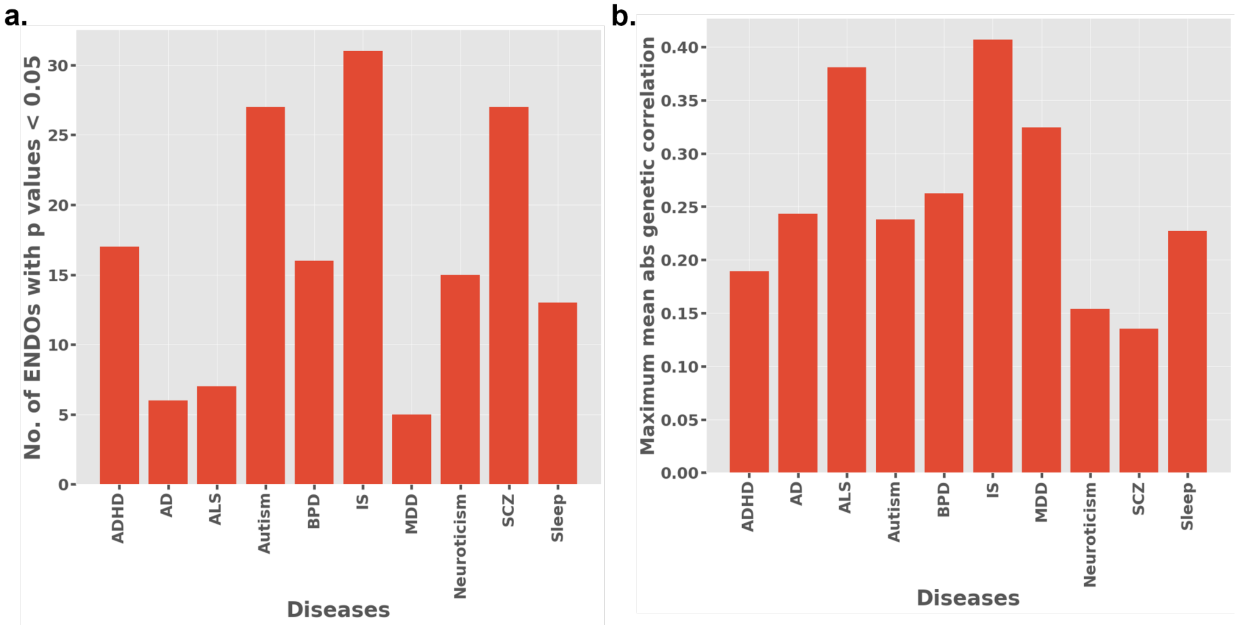


**Supplementary Figure 19.**

a. Number of ENDOs with p-value <0.05 for genetic correlation results for various diseases b. Maximum value of mean absolute genetic correlation for various brain-related conditions. (refer Supplementary Figure 12 for mean abs values for each ENDOs)

ADHD: Attention-deficit/hyperactivity disorder, ALS: Amyotrophic lateral sclerosis, BPD: Bipolar disorder, IS: Ischemic stroke, MDD: Major depressive disorder, Sleep: Sleep disorder

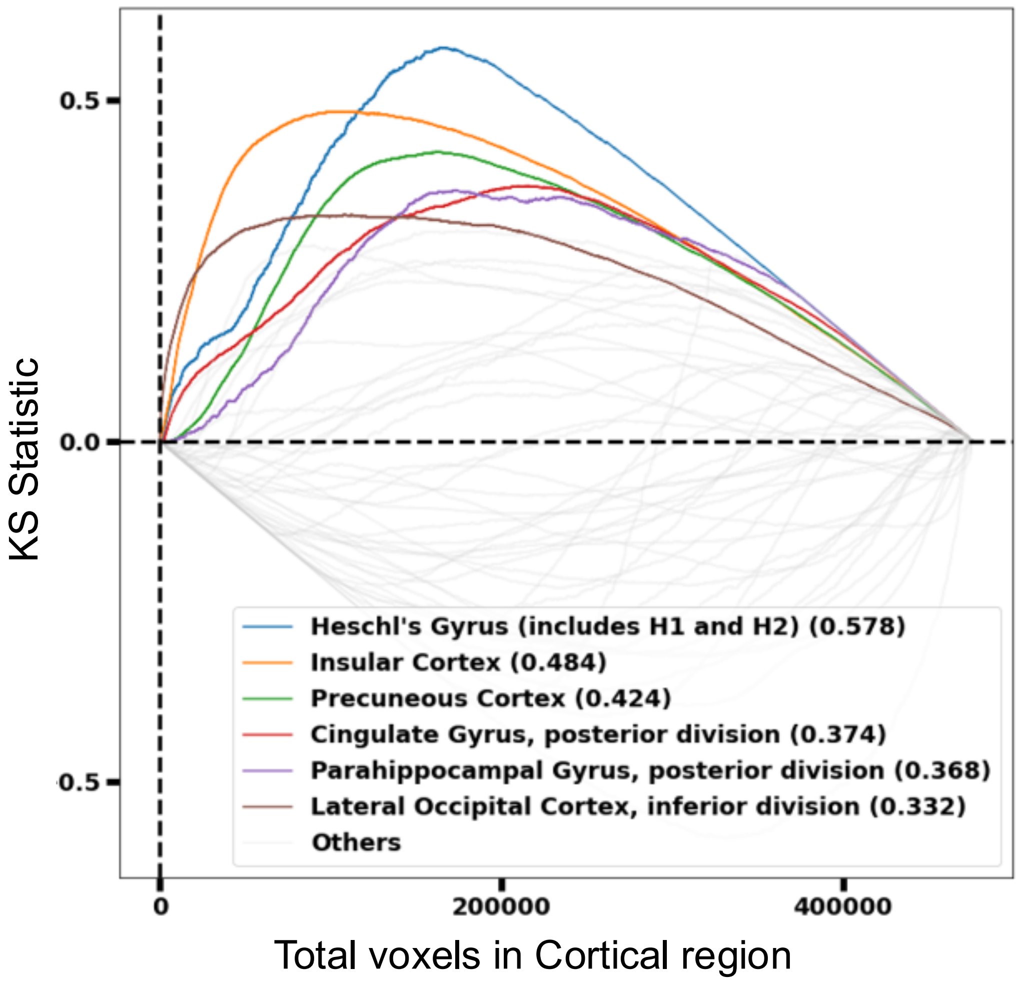


**Supplementary Figure 20. K-S statistics plot for cortical region t-map for ENDO T2:67.** Harvard-Oxford cortical atlas was used to select regions of t-map generated through PerDI for ENDO T2:67. Voxels in the cortical atlas were ranked and K-S statistic plot was generated.

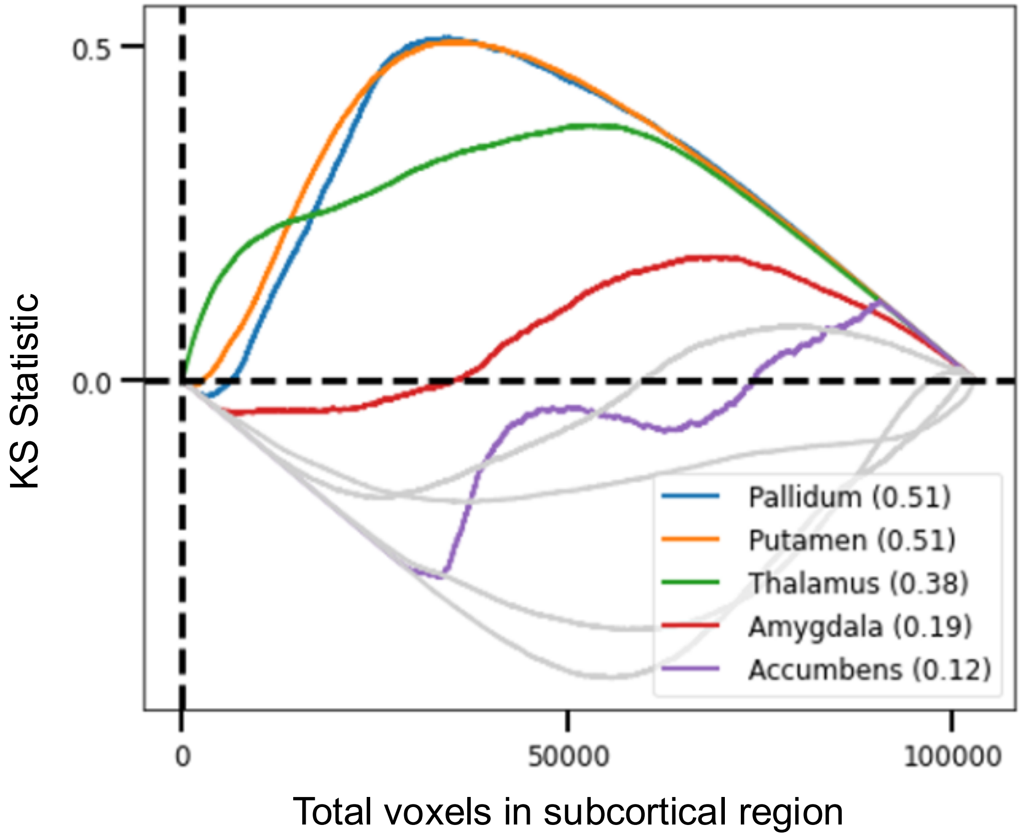


**Supplementary Figure 21. K-S statistics plot for subcortical region t-map for ENDO T2:67.** Harvard-Oxford subcortical atlas was used to select regions of t-map generated through PerDI for ENDO T2:67. Voxels in the subcortical atlas were ranked and K-S statistic plot was generated.

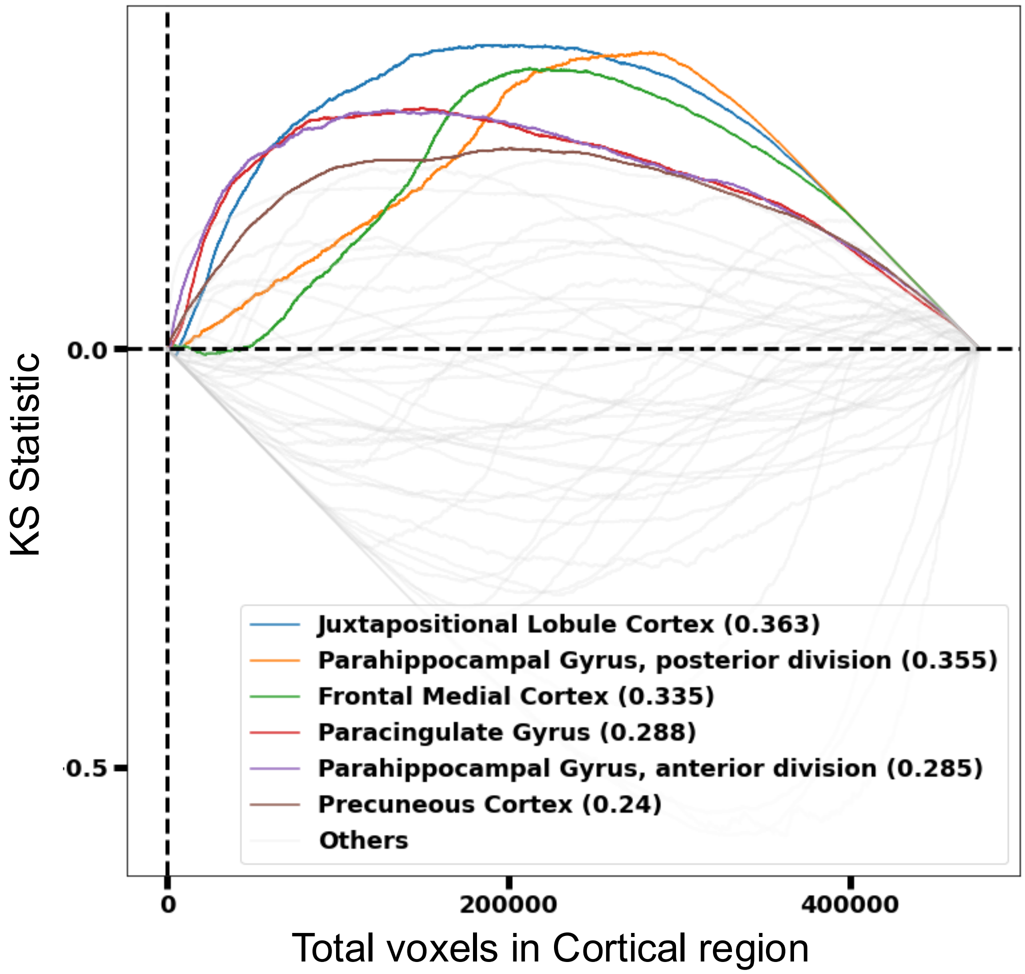


**Supplementary Figure 22. K-S statistics plot for cortical region t-map for ENDO T2:14.** Harvard-Oxford cortical atlas was used to select regions of t-map generated through PerDI for ENDO T2:14. Voxels in the cortical atlas were ranked and K-S statistic plot was generated.

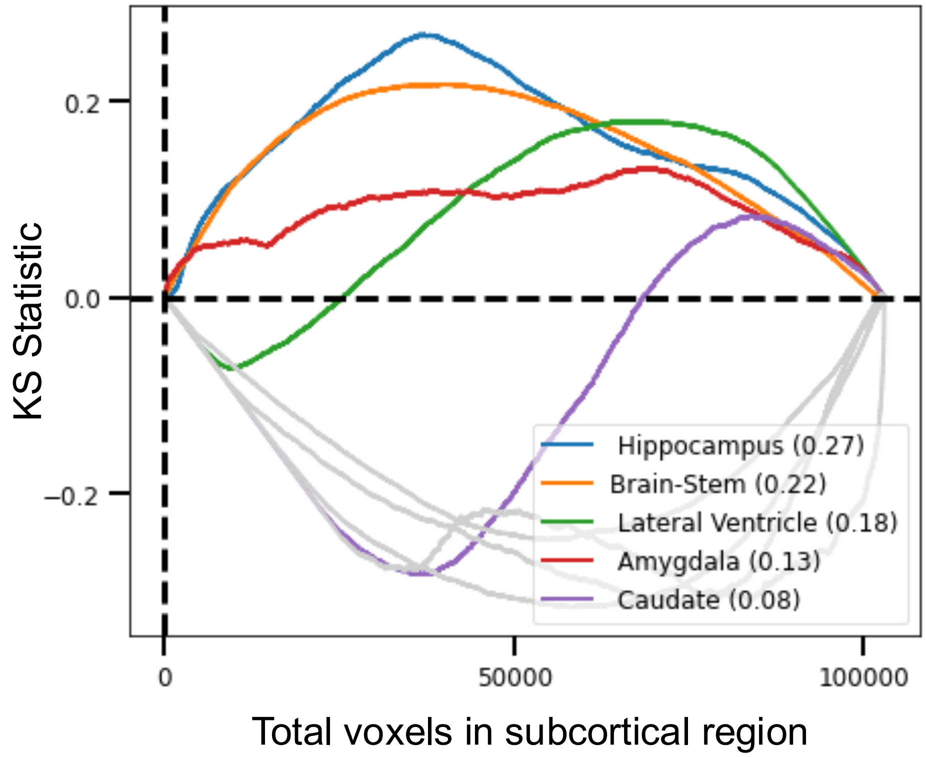


**Supplementary Figure 23. K-S statistics plot for subcortical region t-map for ENDO T2:14.** Harvard-Oxford subcortical atlas was used to select regions of t-map generated through PerDI for ENDO T2:14. Voxels in the subcortical atlas were ranked and K-S statistic plot was generated.

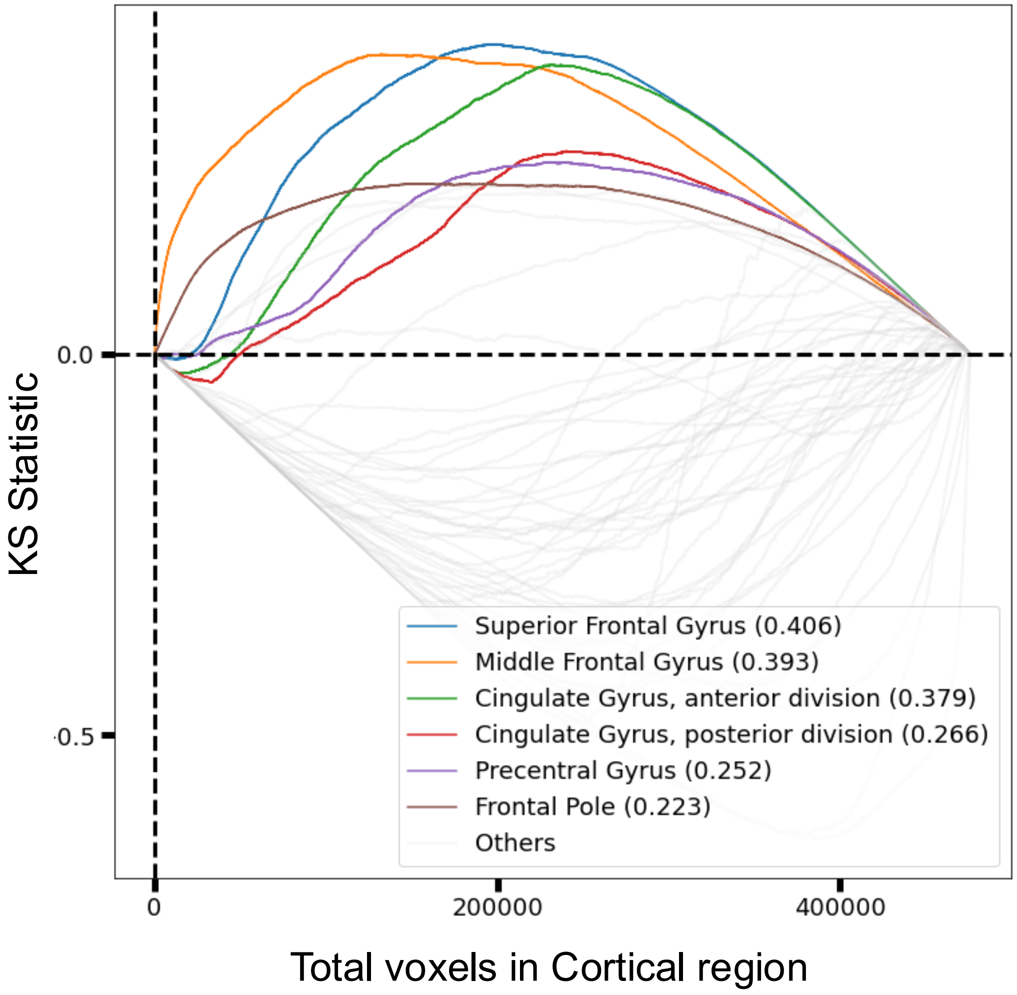


**Supplementary Figure 24. K-S statistics plot for cortical region t-map for ENDO T2:64.** Harvard-Oxford cortical atlas was used to select regions of t-map generated through PerDI for ENDO T2:64. Voxels in the cortical atlas were ranked and K-S statistic plot was generated.

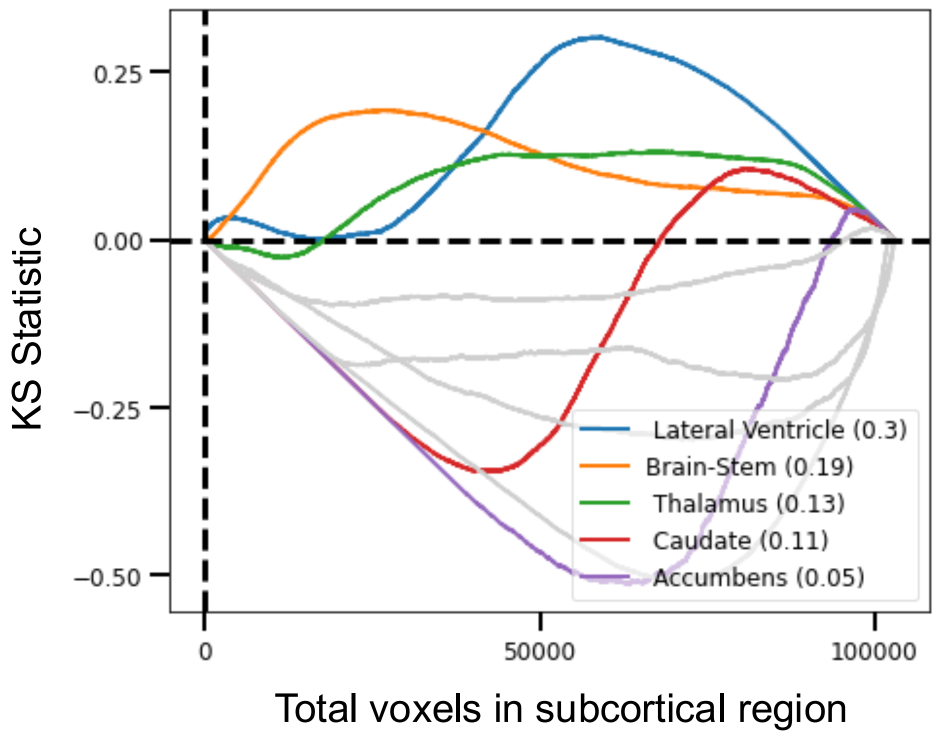


**Supplementary Figure 25. K-S statistics plot for subcortical region t-map for ENDO T2:64.** Harvard-Oxford subcortical atlas was used to select regions of t-map generated through PerDI for ENDO T2:64. Voxels in the subcortical atlas were ranked and K-S statistic plot was generated.

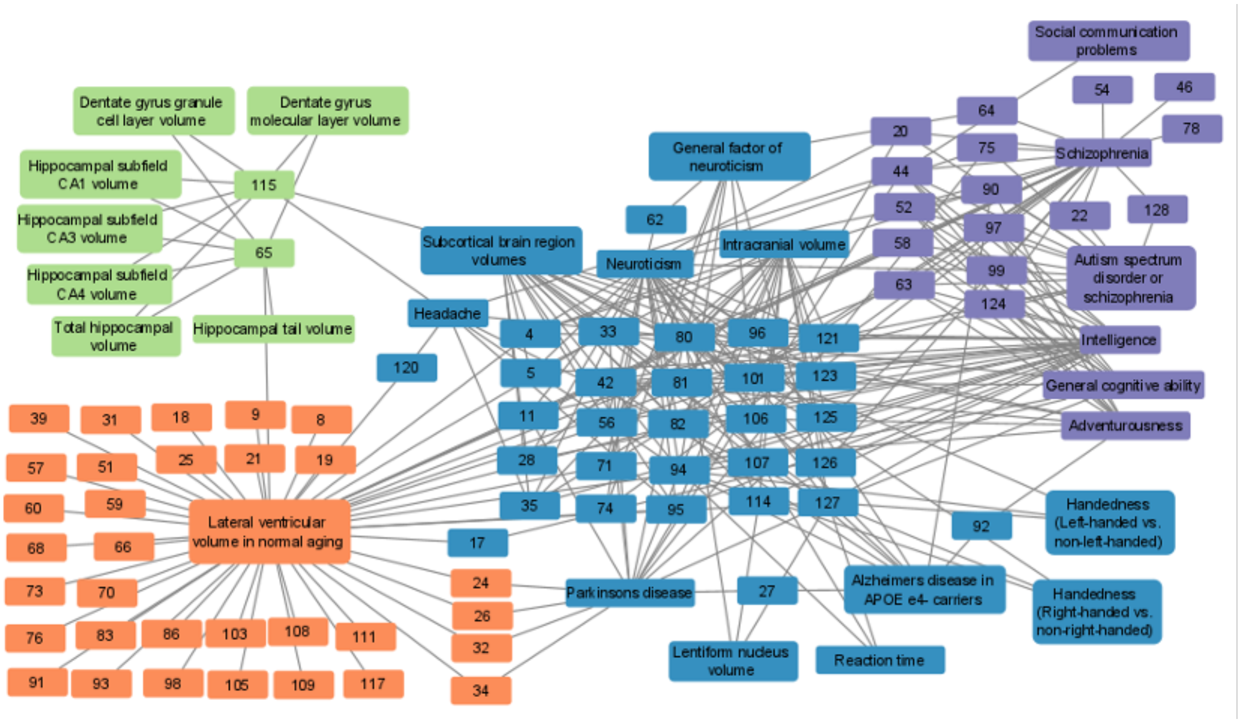


**Supplementary Figure 26. Network of association between ENDOs (numbered nodes) and phenotype discovered in T1.**

Different colors denote different clusters found through Spectral Co-Clustering.

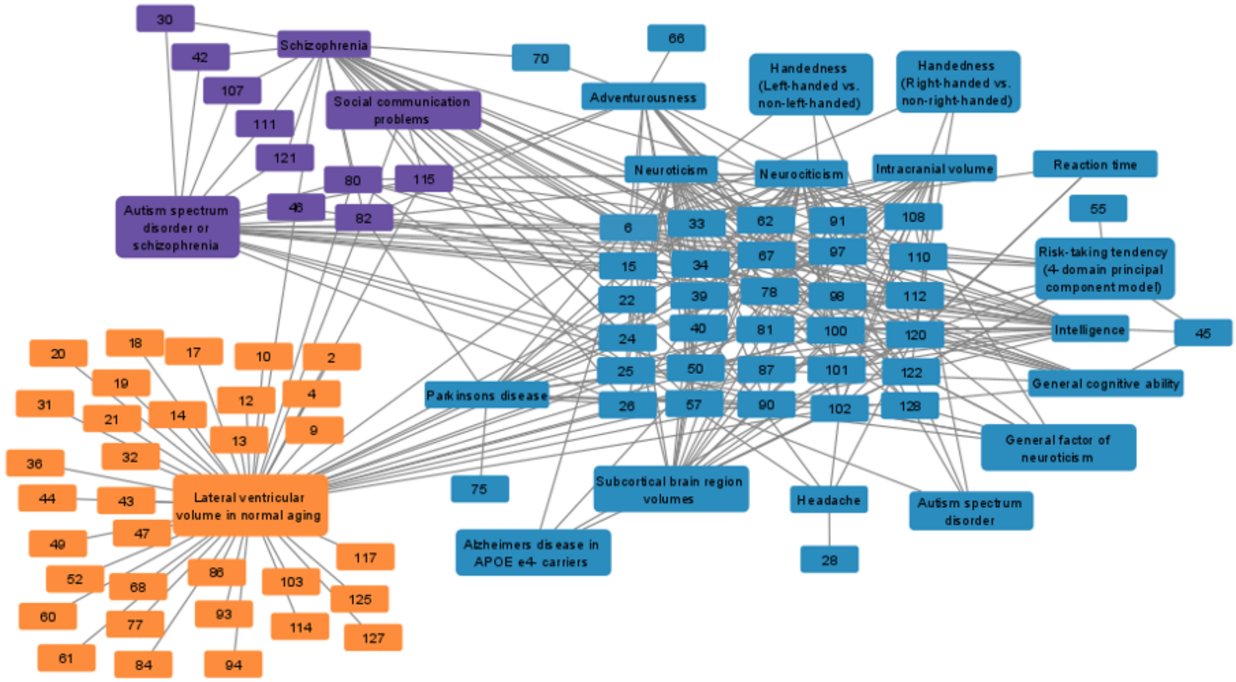


**Supplementary Figure 27. Network of association between ENDOs (numbered nodes) and phenotype discovered in T2.**

Different colors denote different clusters found through Spectral Co-Clustering.

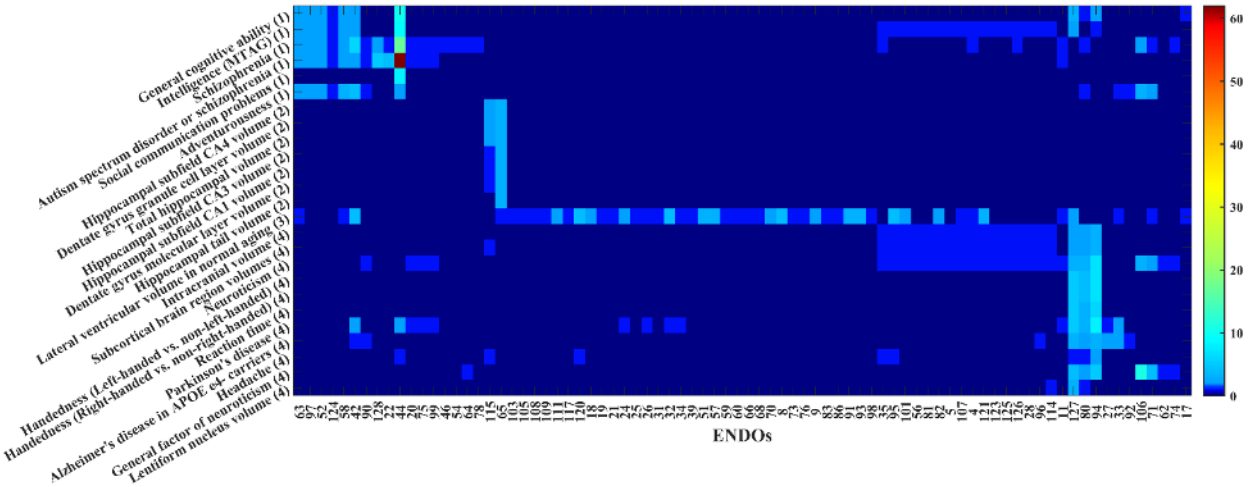


**Supplementary Figure 28. Bi-clustering view of association between ENDOs (numbered nodes) and phenotype discovered in T1.**

Colors denote the size of the shared genes between each ENDO and phenotype.

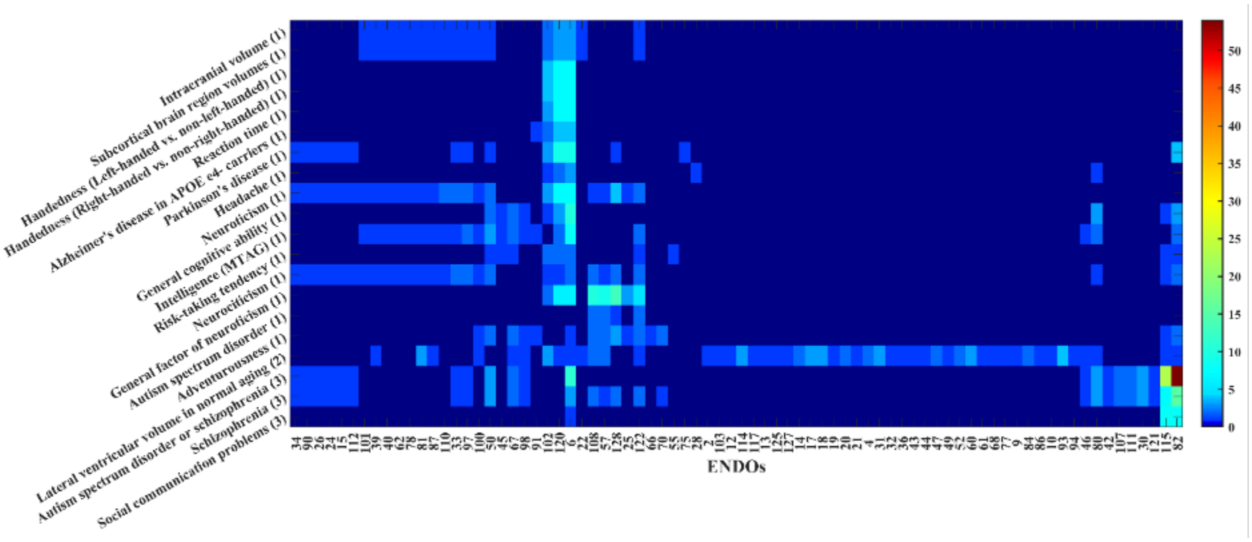


**Supplementary Figure 29. Bi-clustering view of association between ENDOs (numbered nodes) and phenotype discovered in T2.**

Colors denote the size of the shared genes between each ENDO and phenotype.

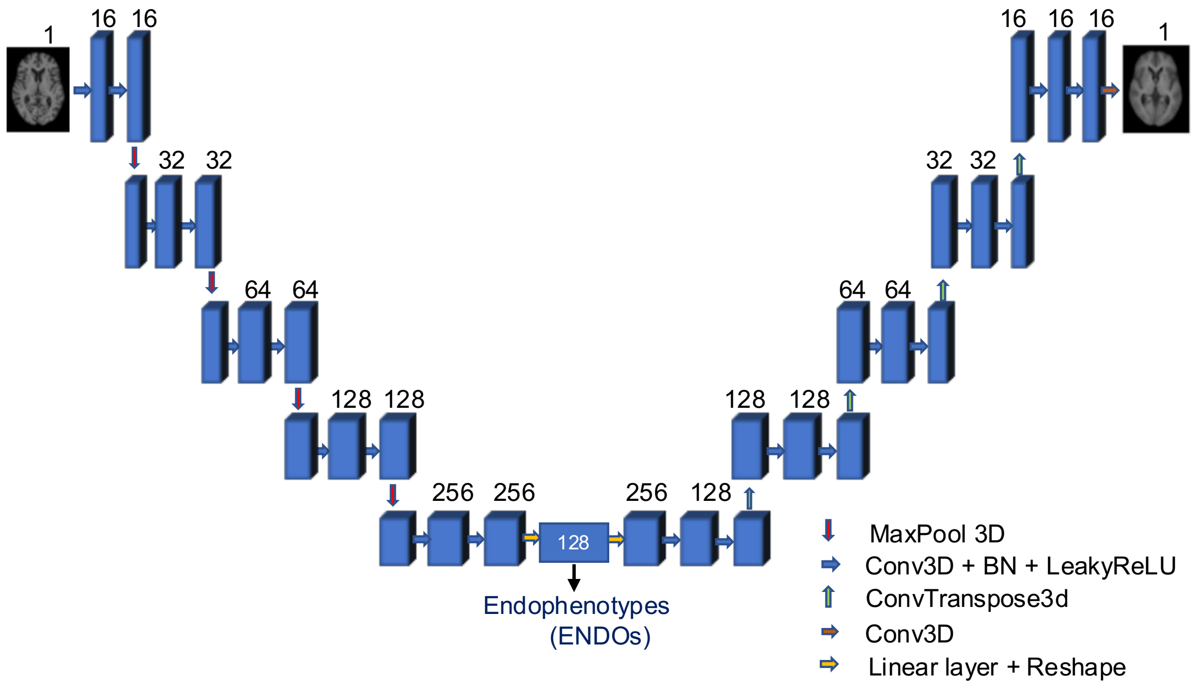


**Supplementary Figure 30. 3D convolutional autoencoder architecture.**
3D convolutional autoencoder comprises four blocks of encoder and four blocks of decoder using 3D convolutional layers. 128-dimensional latent space is used as endophenotypes for GWAS.
