## Supplementary notes for "New phenotype discovery method by unsupervised deep representation learning empowers genetic association studies of brain imaging"

### Supplementary Note 1: Gene-based GWAS catalog analysis using FUMA

Rather than focusing on individual GWAS loci, we used FUMA pipelines to annotate genes and conduct functional enrichment of genes for ENDO GWAS summary statistics. For T2 ENDOs, first, by the positional gene prioritization pipeline (gene assignment pipeline) from FUMA, a total of 353 genes were found (Supplementary Table 10). Using the Gene2Func pipeline of FUMA which performed Gene set enrichment analysis (GSEA), we identified 402 gene sets (adjusted p-value <0.05) using 20260 background genes (protein coding genes) (Supplementary Table 11). To better understand which phenotypes are implicated by our ENDOs, we further analyzed the gene set enriched with various phenotypes of the GWAS Catalog used in FUMA. Out of all the phenotypes, the *Autism spectrum disorder or schizophrenia-related* gene set was the most significantly enriched gene set with an adjusted p-value of $6.74X10^{-38}$. Additionally, gene sets from 19 other brain-related phenotypes from GWAS catalogs were also significantly enriched, including for example, General factor of neuroticism (adjusted p-value = $7.28X10^{-21}$), Handedness (adjusted p-value = $1.52X10^{-10}$), Parkinson's disease (adjusted p-value = $2.54X10^{-7}$), and Alzheimer's disease in APOE e4- carriers (adjusted p-value = $3.93X10^{-4}$).

The same procedure was followed for T1 ENDOs, and 432 genes were found (Supplementary Table 12). Three hundred forty-four gene sets were enriched (Supplementary Table 13). Overall, more brain-related phenotypes were associated with T1 ENDOs than T2 ENDOs (25 vs. 20). Similar to T2 ENDOs, *Autism spectrum disorder or schizophrenia-related* gene sets were the most significant gene sets. But more brain structure-related phenotypes like Hippocampal subfield CA4 volume, Total hippocampal volume, Hippocampal subfield CA3 volume, and Hippocampal subfield CA1 volume are enriched for T1 ENDOs. See **Gene annotation** for more information.

To aid further interpretation of the ENDOs with 25 T1 and 20 T2 brain-related GWAS Catalog phenotypes, we built a bipartite graph of ENDOs and phenotypes whose edges are weighted by the number of shared genes associated with the ENDO and the phenotype by FUMA. Although the bipartite graph is all interconnected (Supplementary Figures 26-27), we identified four robust bi-clusters of ENDOs and brain-associated phenotypes in T1 and three robust clusters in T2 using a mixture of clustering method to determine the most robust number of phenotype clusters across different resolutions (Quantum Clustering[1], see **Bi-Clustering ENDOs and phenotypes )** and bi-clustering of phenotypes and ENDOs (Spectral CoClustering[2], see **Bi-Clustering ENDOs and phenotypes**). Lateral ventricular volume in normal aging formed an independent cluster with multiple ENDOs associated only with it in both T1 and T2 (Supplementary Figures 26-29). Not surprisingly, *Schizophrenia*, *Autism spectrum disorder or schizophrenia* and *Social communication problems* cluster together in both T1 and T2. Interestingly, they cluster with *General cognitive activity, Intelligence* and *adventurousness* in T1 but *Intelligence* and *adventurousness* are in a different cluster in T2 (Supplementary Figures 26-29). Previous studies indicate that alleles for autism overlap broadly with alleles for high intelligence[3]. T1 has a unique cluster that does not appear in T2 and is driven by two ENDOs (65 and 115) and gene sets involved in several hippocampal locations (Dentate gyrus granule cell layer volume and molecular layer volume, Hippocampal subfield tail, CA1, CA3 and CA4 subfields and total Hippocampal volumes).

#### Gene annotation

FUMA[4] was used to annotate our GWAS results functionally from February 26, 2022 till April 15,2022. FUMA has integrated the functional annotation pipeline, including SNP annotation, gene mapping, and gene set enrichment analysis. We used the summary statistics of GWAS as input and identified the prioritized genes through functionally annotated variants. We used the default setting in the FUMA web interface to find positionally prioritized genes within a 10 kb window of functionally associated variants. To assess the functionally enriched phenotypes associated with our ENDOs, we performed gene set enrichment analysis in FUMA with all unique genes identified in our ENDOs.

#### Bi-clustering ENDOs and phenotypes

ENDOs were associated with phenotypes based on the number of shared genes that are mutually associated with them, where genes associated with ENDOs were retrieved from FUMA[1,4] and genes associated with a phenotype were retrieved from GWAS Catalog. In order to bi-cluster the ENDOs and phenotypes, we used a two-step process. The first step was used to assess the number of robust clusters. We applied Quantum Clustering[1] with pre-processing involving top principal components from singular value decomposition and whitening of the data (normalizing to unit vector) across different resolutions, as described in Maignan et al[5]. Quantum clustering receives a resolution parameter (sigma - width of the potential function), ranging from zero to 1. We ran the algorithm with sigma ranging between 0.2 and 0.75, resulting in clusters between 10 with the highest resolution and one with the lowest. The number of clusters that remained the same across the widest set of sigma values was considered the most robust (four for T1, with sigma between 0.29 and 0.56 and three for T2 with sigma between 0.32 and 0.68). We then ran the Spectral Co-Clustering algorithm[2] with the number of robust clusters identified in step one to obtain the bi-clustering of the ENDOs and phenotypes. We used Cytoscape version 3.9.1 to plot the T1 and T2 networks formed by the association between ENDOs and phenotypes and color them based on the aforementioned bi-clusters.

### Supplementary Note 2: Learning rate for deep learning training

We used Adam optimizer with initial learning rate (lr) of 0.0005248074602497723 for T1 and 0.0003019951720402019 for T2 was used with a batch size of 62. Learning rate finder (LR finder) function of the Pytorch Lightning framework was used to obtain the initial optimal lr. LR finder increases lr after each mini-batch and plots a lr vs loss plot. The plot is then used to select optimum initial lr. Learning rate was reduced by half when validation set MSE loss plateaued for four epochs with a lower cap of lr/1000.
